# CARDIAC-FM: A Generalizable Multimodal Foundation Model Integrating ECG and Cardiac MRI

**DOI:** 10.64898/2026.03.16.26348526

**Authors:** Fumin Li, Siting Li, Bojun Chen, Yuhan Qian, Jennifer A Brody, Vidhushei Yogeswaran, Kerri L Wiggins, Colleen M Sitlani, Joshua C Bis, Ali Shojaie, W T Longstreth, Bruce M Psaty, Geoffrey H Tison, Simon Du, James S Floyd, Ting Ye

## Abstract

Atrial fibrillation and heart failure impose substantial health burdens worldwide, yet accurate and generalizable risk prediction remains challenging. Here we developed CARDIAC-FM, a multimodal foundation model that integrates 12-lead electrocardiography (ECG) and cardiac magnetic resonance imaging (cardiac MRI) through self-supervised representation learning and cross-modal contrastive alignment. CARDIAC-FM introduces a self-supervised spatiotemporal, multi-view masked autoencoder for cardiac MRI. Developed on 57,609 paired ECG-cardiac MRI samples from UK Biobank, CARDIAC-FM improved prediction of incident atrial fibrillation and heart failure over contemporary ECG AI models and generalized zero-shot to two external cohorts, the Cardiovascular Health Study and the Multi-Ethnic Study of Atherosclerosis. Combining its ECG representation with established clinical risk scores further improved discrimination across cohorts, supporting complementary prognostic information from ECG and traditional risk factors. Beyond atrial fibrillation and heart failure, the learned representation transferred to continuous cardiac MRI phenotype prediction, time-to-event modelling and prediction of additional cardiovascular outcomes with limited fine-tuning, including myocardial infarction, ischaemic stroke, cardiovascular death and all-cause mortality. Although pre-trained with paired ECG and cardiac MRI, the model can be deployed using ECG alone, with additional predictive gains when cardiac MRI is available. These findings demonstrate the promise of multimodal self-supervised learning for generalizable cardiovascular risk prediction.

## Introduction

Atrial fibrillation and heart failure cause substantial cardiovascular morbidity and mortality worldwide, and their prevalence is projected to rise substantially over the coming decades^1–3^. Early identification of individuals at elevated risk is essential for targeted prevention, clinical decision-making, risk stratification across both clinical practice and trial design. However, existing risk prediction models based on traditional cardiovascular risk factors show varying discrimination and limited generalizability across populations^4–6^.

Deep learning applied to the resting 12-lead electrocardiogram (ECG) has shown promise for cardiovascular risk assessment. Convolutional neural networks trained on ECGs can detect subclinical cardiovascular conditions – including structural heart disease^7^, left atrial abnormalities^8,9^, ventricular dysfunction^10^ – and identify existing or near-term atrial fibrillation^11–13^ and predict other cardiovascular outcomes^14–16^. More recently, ECG foundation models trained with self-supervised learning on unlabeled data have further improved representation learning from electrocardiographic signals^17–19^. In parallel, deep learning applied to cardiac magnetic resonance imaging (MRI) has enabled automated cardiovascular disease screening and diagnosis^20–23^.

Despite these advances, current artificial intelligence (AI) models share several limitations that constrain clinical translation. Most models rely on a single modality, even though complementary data sources capture distinct and synergistic aspects of cardiac pathophysiology. Models are often developed in highly selected clinical cohorts, which limits their utility for risk prediction and population-based screening in apparently healthy individuals. Most models are also task-specific, requiring separate training for each outcome of interest. Finally, few have demonstrated robust generalizability across populations with different demographic characteristics, risk profiles, and healthcare contexts.

Recent multimodal approaches have sought to integrate ECG with additional data modalities^23–25^ or combine imaging with clinical text^26^, but have largely focused on identifying prevalent disease rather than predicting future cardiovascular events. The most directly comparable ECG–MRI approach used convolutional neural network (CNN)-based autoencoders jointly trained with reconstruction and cross-modal objectives, with fixed representations for downstream prediction^24^. More broadly, whether multimodal pretraining can enable prospective cardiovascular risk prediction, adaptation across distinct downstream tasks and generalization to independent populations remains incompletely understood.

Here, we introduce CARDIAC-FM (Contrastive Alignment for Risk Detection In Cardiovascular Foundation Model), a multimodal foundation model that integrates large-scale 12-lead ECG and cardiac MRI data from UK Biobank. CARDIAC-FM uses a two-stage training framework that combines modality-specific self-supervised pretraining, transformer-based encoders, and cross-modal alignment with task-specific fine-tuning. In the first stage, modality-specific ECG and cardiac MRI encoders are aligned using paired samples through contrastive learning in a shared latent space. For cardiac MRI, we develop a self-supervised spatiotemporal, multi-view masked autoencoder, trained from scratch on raw cine MRI without phenotype or outcome supervision. The ECG encoder is initialized from a previously pretrained ECG foundation model (ECG-FM^18^). Cross-modal alignment enables the ECG representation to incorporate information captured by cardiac MRI. In the second stage, a lightweight prediction head is added and fine-tuned for downstream outcome prediction, allowing efficient adaptation to various clinical endpoints without training a separate model from scratch for each outcome. This framework is designed to leverage large-scale unlabeled multimodal data while reducing dependence on outcome-specific labeled data and resource-intensive training of separate models for each outcome.

At inference, CARDIAC-FM can operate using ECG alone or ECG with cardiac MRI and can be further combined with established clinical risk scores, accommodating differences in data availability across clinical settings. We evaluated CARDIAC-FM across distinct downstream tasks, including binary incident cardiovascular outcome prediction, continuous cardiac MRI phenotype prediction and time-to-event modelling. We assessed incident atrial fibrillation and heart failure in UK Biobank and evaluated zero-shot generalizability in two independent, multi-site prospective US cohorts—the Cardiovascular Health Study (CHS)^27^ and the Multi-Ethnic Study of Atherosclerosis (MESA)^28^. We further evaluated transfer to additional cardiovascular outcomes, including myocardial infarction, ischaemic stroke, cardiovascular death and all-cause mortality, using limited-data fine-tuning to reflect the scarcity of labelled outcomes in external cohorts. CARDIAC-FM was benchmarked against contemporary ECG AI models using common training and evaluation protocols, and its ECG representation was evaluated for added predictive value beyond established clinical risk scores. Together, these analyses evaluate whether multimodal pretraining yields representations that generalize across populations and transfer across cardiovascular outcomes and prediction tasks. To our knowledge, CARDIAC-FM is the first ECG–cardiac MRI multimodal foundation model for prospective cardiovascular risk prediction externally validated across multiple independent cohorts. We publicly release the model code and pretrained weights to facilitate reproducibility and further research.

## Results

### UK Biobank Data and Participant Characteristics

At the time of analysis, resting 12-lead ECG and cardiac MRI data were available from 71,144 UK Biobank participants who had attended the baseline imaging visit. Participants had a mean age of 65.6 years (s.d. 7.8); 51.4% were female, and a total of 6,182 (8.7%) returned for repeat imaging, with a median inter-scan interval of 3.0 years (interquartile range 2.2-3.4 years), yielding 77,326 imaging visits (Supplementary Tables 1-2). ECG and MRI were acquired on the same day at each visit.

Among all the imaging visits, 70,424 visits had evaluable MRI and 67,661 had evaluable ECG data. Most visits (80.3%) had both modalities available, 7.2% had ECG only, and 10.7% had MRI only (Supplementary Table 1). After applying pre-specified quality control criteria (Methods), a total of 57,609 visits with paired, high-quality ECG and MRI data were retained for analysis. Participants with prevalent atrial fibrillation or heart failure at baseline were retained for pretraining but excluded from outcome prediction analyses. We randomly partitioned participants into training (60%), validation (5%), and test (35%) sets at the participant level, ensuring that all visits from the same individual were assigned to a single partition.

### Overview of Multimodal AI model

We developed CARDIAC-FM as a multimodal self-supervised framework that integrates raw ECG and cardiac MRI inputs (Fig. 1). The cardiac MRI encoder is a new spatiotemporal, multi-view masked autoencoder, pretrained from scratch on raw cine MRI without phenotype or outcome supervision. The encoder jointly processes short-axis, four-chamber and two-chamber views using spatiotemporal masking and a Vision Transformer^29^ architecture. The resulting cardiac MRI representation is aligned with the pretrained ECG representation in a shared latent space using a symmetric InfoNCE contrastive loss^30^. The model was trained on 34,596 samples with paired ECG and MRI data. For downstream outcome prediction, we excluded participants with prevalent disease at baseline, added a prediction layer to the learned representations, and fine-tuned the model for 20 epochs using supervised learning. CARDIAC-FM supports two modality configurations, ECG-only and ECG+MRI; either prediction can additionally be combined with a clinical risk score through late fusion. Full methodological details are provided in Methods.

**Figure 1:**
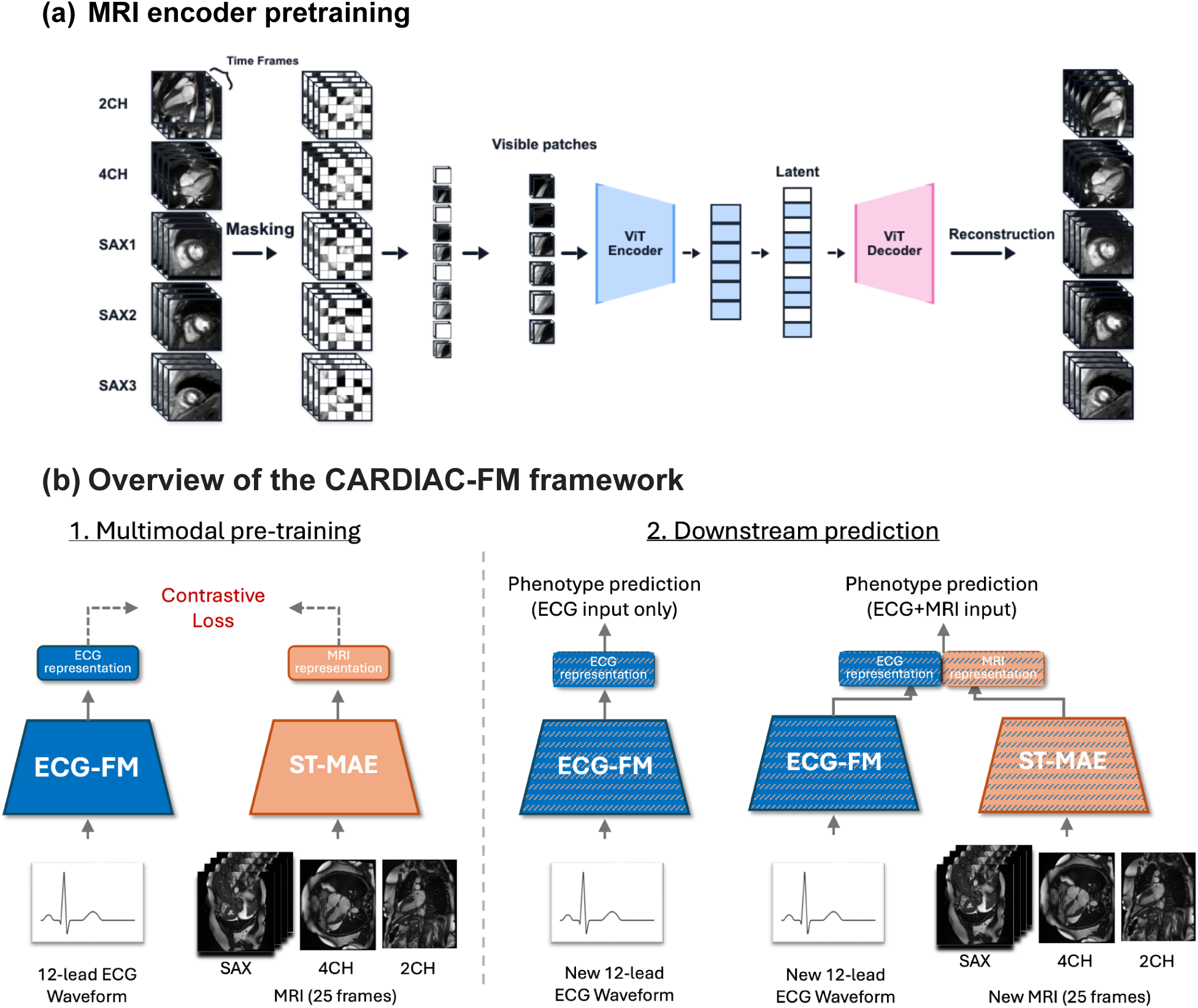
CARDIAC-FM architecture and study cohorts. (a) Self-supervised cardiac-MRI pretraining. Multi-view cine MRI sequences, including short-axis (SAX), four-chamber (4CH), and two-chamber (2CH) views, are divided into spatiotemporal patches and randomly masked. Visible patches are encoded by a Vision Transformer (ViT) and passed to a decoder to reconstruct the original MRI inputs. (b) Multimodal CARDIAC-FM framework. ECG waveforms are processed by the ECG Foundation Model (ECG-FM) encoder and MRI by the pretrained spatiotemporal multi-view masked autoencoder (ST-MAE). Their representations are aligned using symmetric contrastive loss. For downstream prediction, the pretrained ECG encoder can be used independently or combined with the MRI encoder; both configurations can also incorporate established clinical risk scores.

We compared CARDIAC-FM with established clinical risk scores (CHARGE-AF for atrial fibrillation and PREVENT-HF for heart failure)^6,31^ and four contemporary ECG AI models (ECG-FM, ECGFounder, DeepECG-SL, and DeepECG-SSL)^18,32,33^. All ECG models were initialized from their respective pretrained weights and fine-tuned using the same protocol as CARDIAC-FM. We evaluated CARDIAC-FM using ECG only or combined ECG and cardiac MRI, and further evaluated its performance when combined with the corresponding clinical risk score through late fusion. Performance was evaluated using the area under the receiver operating characteristic curve (AUROC) and area under the precision–recall curve (AUPRC). We obtained 95% confidence intervals (CIs) and assessed pairwise differences between models by bootstrapping the held-out test set.

### Fine-Tuning for Cardiac MRI Feature Prediction

We fine-tuned CARDIAC-FM with ECG-only input to predict continuous MRI-derived cardiac structural and functional measures: left ventricular ejection fraction (LVEF), left ventricular mass (LVM), left ventricular end-diastolic volume (LVEDV), left ventricular end-systolic volume (LVESV), left atrial emptying fraction (LAEF), left atrial minimum volume (LAVmin), and left atrial maximum volume (LAVmax). ECG-predicted values were strongly correlated with direct imaging measures across all features (Pearson r=0.45-0.79; *R*^2^= 0.20-0.60; Supplementary Fig. 1; Supplementary Table 20). Correlations were generally higher for structural measures (LVM, *r* = 0.79; LVEDV, *r* = 0.72; LVESV, *r* = 0.71; LAVmin, *r* = 0.56) than for functional indices (LVEF, *r* = 0.49; LAEF, *r* = 0.45), with LAVmax showing intermediate performance (*r* = 0.53). These findings demonstrate that ECG representations learned through multimodal pretraining capture structural and functional cardiac information acquired with MRI, enabling imputation of imaging-derived features when cardiac MRI is unavailable.

### Fine-Tuning for Downstream Clinical Outcome Prediction

We evaluated CARDIAC-FM for 5-year prediction of incident atrial fibrillation and heart failure in the held-out UK Biobank test set. Analyses were restricted to participants with both ECG and cardiac MRI available, enabling direct comparison across input configurations (outcome rates in Supplementary Table 2; Fig. 2 and Supplementary Table 5). CARDIAC-FM was evaluated using ECG only or combined ECG and cardiac MRI, and was compared with four contemporary ECG AI models (ECG-FM, ECGFounder, DeepECG-SL and DeepECG-SSL) and established clinical risk scores (CHARGE-AF for atrial fibrillation and PREVENT-HF for heart failure).

**Figure 2:**
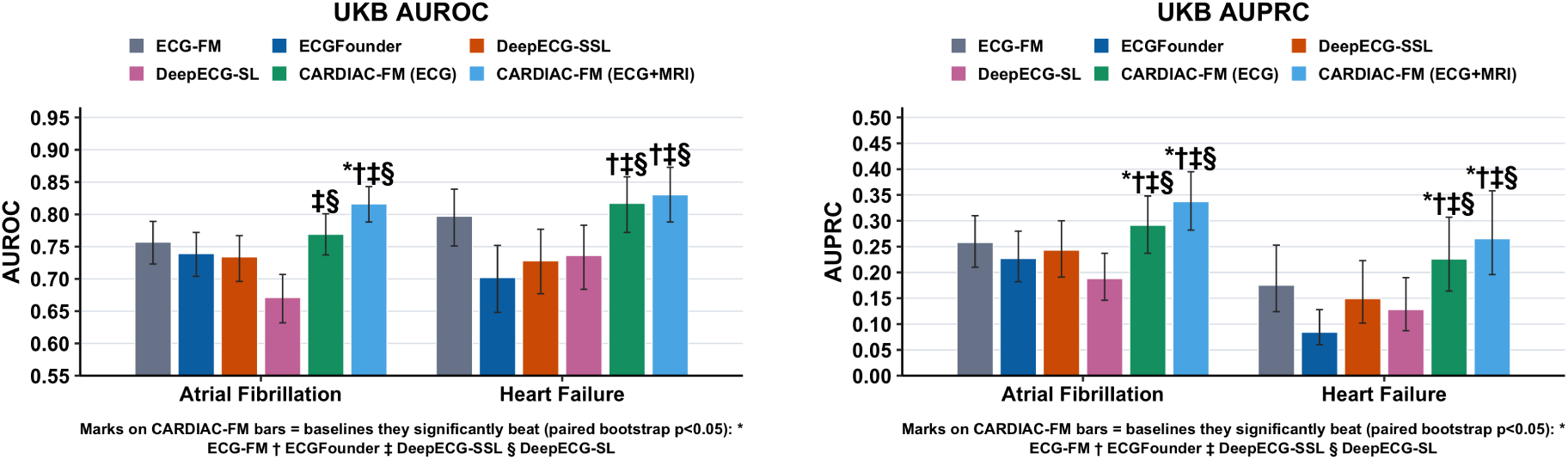
Prediction of incident atrial fibrillation and heart failure in the UK Biobank. AUROC (left) and AUPRC (right) for 5-year prediction of incident atrial fibrillation and heart failure in the held-out test set. Models include ECG-FM, ECGFounder, DeepECG-SSL, DeepECG-SL, CARDIAC-FM(ECG) and CARDIAC-FM(ECG+MRI). Error bars indicate 95% bootstrap confidence intervals. Symbols indicate statistically significant pairwise differences between CARDIAC-FM and comparator ECG AI models based on bootstrap (P < 0.05).

Multimodal pretraining improved prediction even when only ECG was available at inference. For atrial fibrillation, CARDIAC-FM (ECG only) achieved an AUROC of 0.769 (95% CI, 0.737–0.801) and an AUPRC of 0.291 (0.237–0.348). It significantly outperformed two of four contemporary ECG AI models in AUROC and all four in AUPRC (P < 0.05). For heart failure, CARDIAC-FM (ECG only) achieved an AUROC of 0.817 (0.772–0.858) and an AUPRC of 0.226 (0.164–0.307), significantly outperforming three of four ECG AI models in AUROC and all four in AUPRC (P < 0.05).

Adding cardiac MRI at inference further improved performance. For atrial fibrillation, CARDIAC-FM (ECG+MRI) achieved an AUROC of 0.816 (95% CI, 0.788–0.843) and an AUPRC of 0.337 (0.282–0.395), compared with 0.769 and 0.291, respectively, using ECG alone. For heart failure, CARDIAC-FM (ECG+MRI) achieved an AUROC of 0.830 (0.788–0.873) and an AUPRC of 0.265 (0.196–0.358), compared with 0.817 and 0.226 using ECG alone. CARDIAC-FM (ECG+MRI) significantly outperformed all four for atrial fibrillation, and three of four in AUROC (all four in AUPRC) for heart failure. Combining CARDIAC-FM with established clinical risk scores further improved prediction in UK Biobank (Fig. 4). Together, these findings demonstrate that multimodal pretraining improves ECG-based prediction and that complementary imaging and clinical information can provide additional predictive gains.

To assess performance across clinically relevant subgroups, we evaluated CARDIAC-FM stratified by age (<65 versus ≥65 years) and sex (Supplementary Fig. 4 and Supplementary Table 13). Performance was broadly consistent across age and sex subgroups for both atrial fibrillation and heart failure.

### Time-to-Event Modelling of Cardiovascular Outcomes

We next evaluated whether the pretrained representation can be fine-tuned for time-to-event modelling, accounting for censoring (Supplementary Table 27). In UK Biobank, CARDIAC-FM (ECG) achieved Harrell’s C-indices of 0.740 (95% CI, 0.710–0.768) for atrial fibrillation and 0.786 (0.751–0.822) for heart failure. CARDIAC-FM significantly outperformed three of four contemporary ECG AI models for atrial fibrillation and all four for heart failure (P < 0.05). These findings extend the performance of CARDIAC-FM beyond fixed-horizon classification to censored time-to-event prediction.

### Zero-Shot External Validation in CHS and MESA Cohorts

We externally evaluated CARDIAC-FM in two independent US prospective cohort studies: the Cardiovascular Health Study (CHS) and the Multi-Ethnic Study of Atherosclerosis (MESA). CHS enrolled 5,888 adults aged 65 years or older in 1989-1990 and 1992-1993, including predominantly White participants and a smaller proportion of African American individuals, with a mean age of 73 years at baseline. MESA enrolled 6,814 adults aged 45-84 years from four racial and ethnic groups (White, African American, Hispanic, and Chinese American) in 2000-2002, all free of clinical cardiovascular disease at enrollment, with a mean age of 62 years (Supplementary Tables 3 and 4). Both cohorts differed from UK Biobank imaging participants (mean age 65 years) in age distribution and racial and ethnic composition but provided longitudinal surveillance for atrial fibrillation events and central physician adjudication of heart failure, enabling robust assessment of generalizability. In CHS, participants underwent standardized resting 12-lead ECGs at the baseline examination, whereas in MESA participants underwent both ECG and MRI at baseline. We evaluated CARDIAC-FM (ECG only) and CARDIAC-FM (ECG + Risk Score) in both external cohorts (Fig. 3; Supplementary Tables 7 and 9).

**Figure 3:**
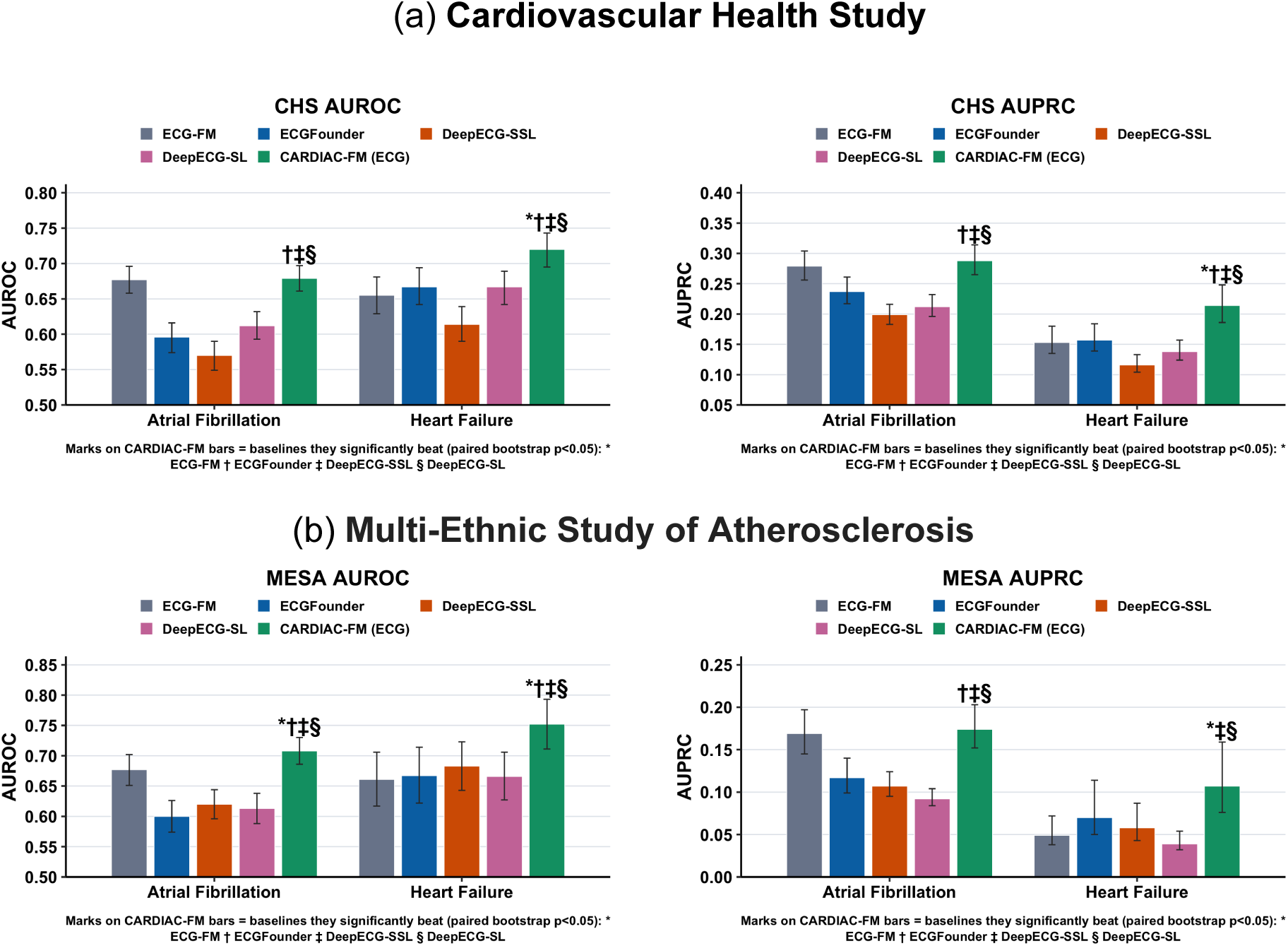
External validation of CARDIAC-FM in (a) the Cardiovascular Health Study and (b) the Multi-Ethnic Study of Atherosclerosis. AUROC (left) and AUPRC (right) for zero-shot 5-year prediction of incident atrial fibrillation and heart failure. Models pretrained in UK Biobank were applied directly to CHS and MESA without cohort-specific fine-tuning. Models include ECG-FM, ECGFounder, DeepECG-SSL, DeepECG-SL and CARDIAC-FM (ECG). Error bars indicate 95% bootstrap confidence intervals. Symbols above CARDIAC-FM bars indicate statistically significant pairwise differences from comparator ECG AI models based on bootstrap (P < 0.05).

In *zero-shot* evaluation, models trained in UK Biobank were applied directly to CHS and MESA without cohort-specific fine-tuning. CARDIAC-FM generalized across both external cohorts, significantly outperforming three of four contemporary ECG AI comparators for both atrial fibrillation and heart failure in AUROC and AUPRC in both CHS and MESA (P < 0.05; Fig. 3 and Supplementary Tables 7 and 9). Combining CARDIAC-FM with established clinical risk scores further improved prediction (Fig. 4 and Supplementary Tables 6, 8 and 10). For heart failure, AUROC increased from 0.751 to 0.792 in CHS and from 0.789 to 0.826 in MESA, with significant improvements in both AUROC and AUPRC in both cohorts (P < 0.05). For atrial fibrillation, AUROC increased from 0.699 to 0.723 in CHS (P < 0.05) but not in MESA (0.775 versus 0.768), whereas AUPRC improved significantly in both cohorts. Together, these findings support zero-shot generalizability of the multimodally pretrained ECG representation and its complementary predictive value beyond established clinical risk factors. Performance in CHS and MESA was also examined within age and sex subgroups (Supplementary Figs. 5 and 6; Supplementary Tables 14 and 15).

**Figure 4:**
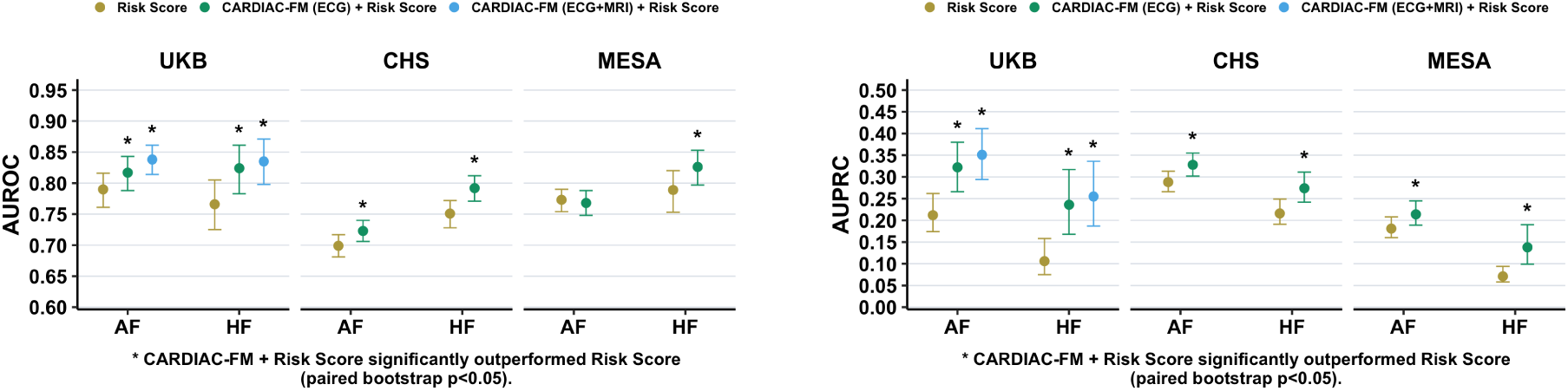
CARDIAC-FM + Risk Score vs Risk Score for prediction of incident atrial fibrillation and heart failure. AUROC (left) and AUPRC (right) for 5-year prediction of incident atrial fibrillation and heart failure in UK Biobank, CHS, and MESA. Models compared include clinical risk scores alone, CARDIAC-FM(ECG)+Risk Score, and CARDIAC-FM(ECG+MRI)+Risk Score where cardiac MRI was available. Error bars indicate 95% bootstrap confidence intervals. CARDIAC-FM combined with clinical risk scores generally improved predictive performance over risk scores alone.

CARDIAC-FM also provided robust risk stratification in both external cohorts (Fig. 5; Supplementary Tables 16). After adjustment for traditional cardiovascular risk factors, the highest versus lowest CARDIAC-FM (ECG + Risk Score) tertile was associated with heart failure hazard ratios (HRs) of 2.74 (95% CI, 2.28–3.30) in CHS and 4.87 (3.14–7.57) in MESA, with particularly strong associations for heart failure with reduced ejection fraction (HFrEF; 4.40 [3.08–6.29] and 7.77 [4.08–14.80], respectively). CARDIAC-FM also retained significant adjusted stratification for atrial fibrillation and heart failure with preserved ejection fraction (HFpEF) in both cohorts. Incremental discrimination analyses similarly favored CARDIAC-FM, with generally positive integrated discrimination improvement (IDI) relative to contemporary ECG AI models and when added to established risk scores (Supplementary Tables 23-24). Absolute risk calibration varied across cohorts but improved substantially after cross-fitted Platt recalibration, with calibration intercepts approaching 0, slopes approaching 1 and generally lower Brier scores; after recalibration, decision-curve analyses generally showed greater net benefit for CARDIAC-FM combined with clinical risk scores than for clinical risk scores alone across clinically relevant thresholds (Supplementary Figs. 7-9 and Supplementary Table 25).

**Figure 5:**
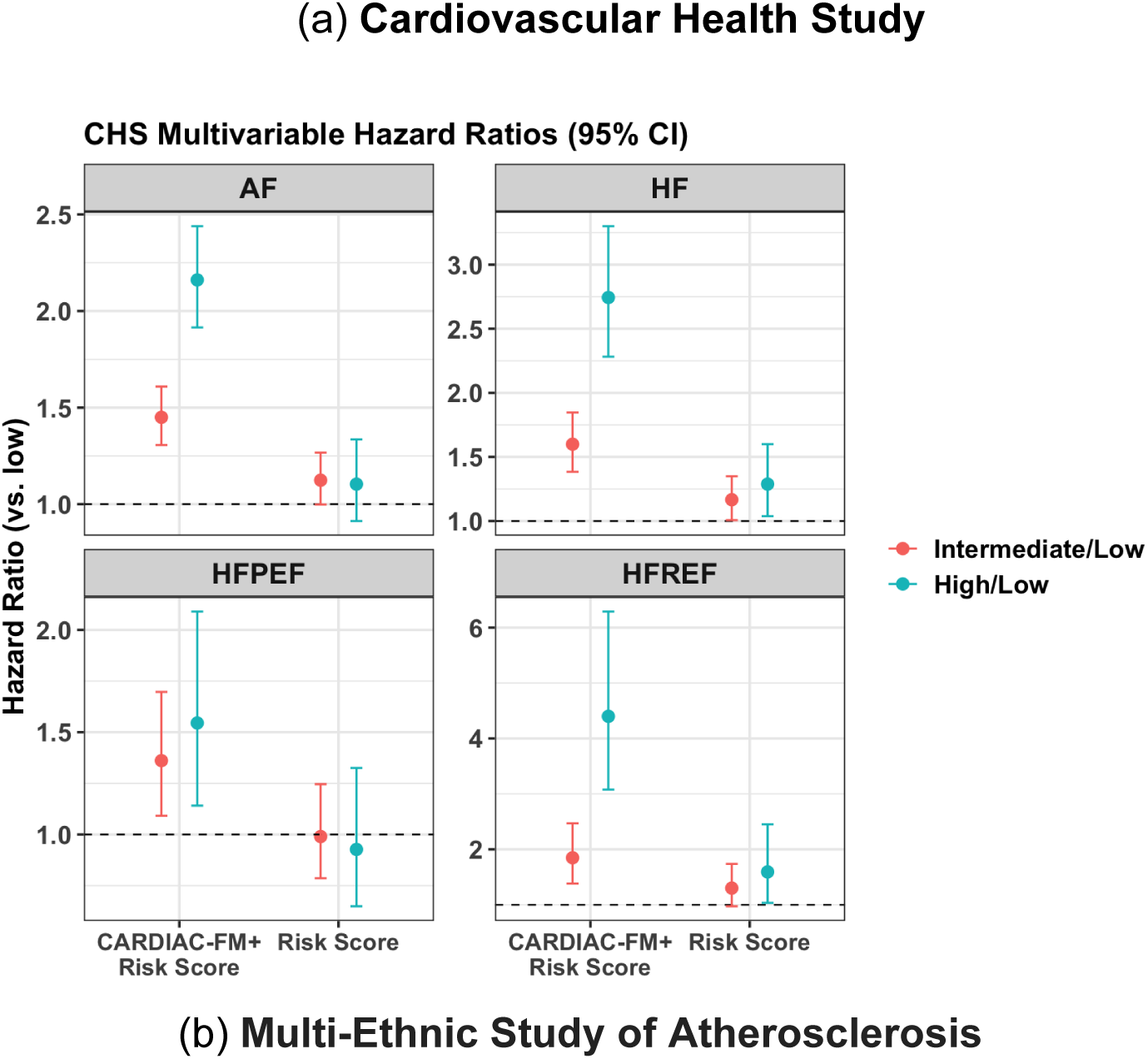
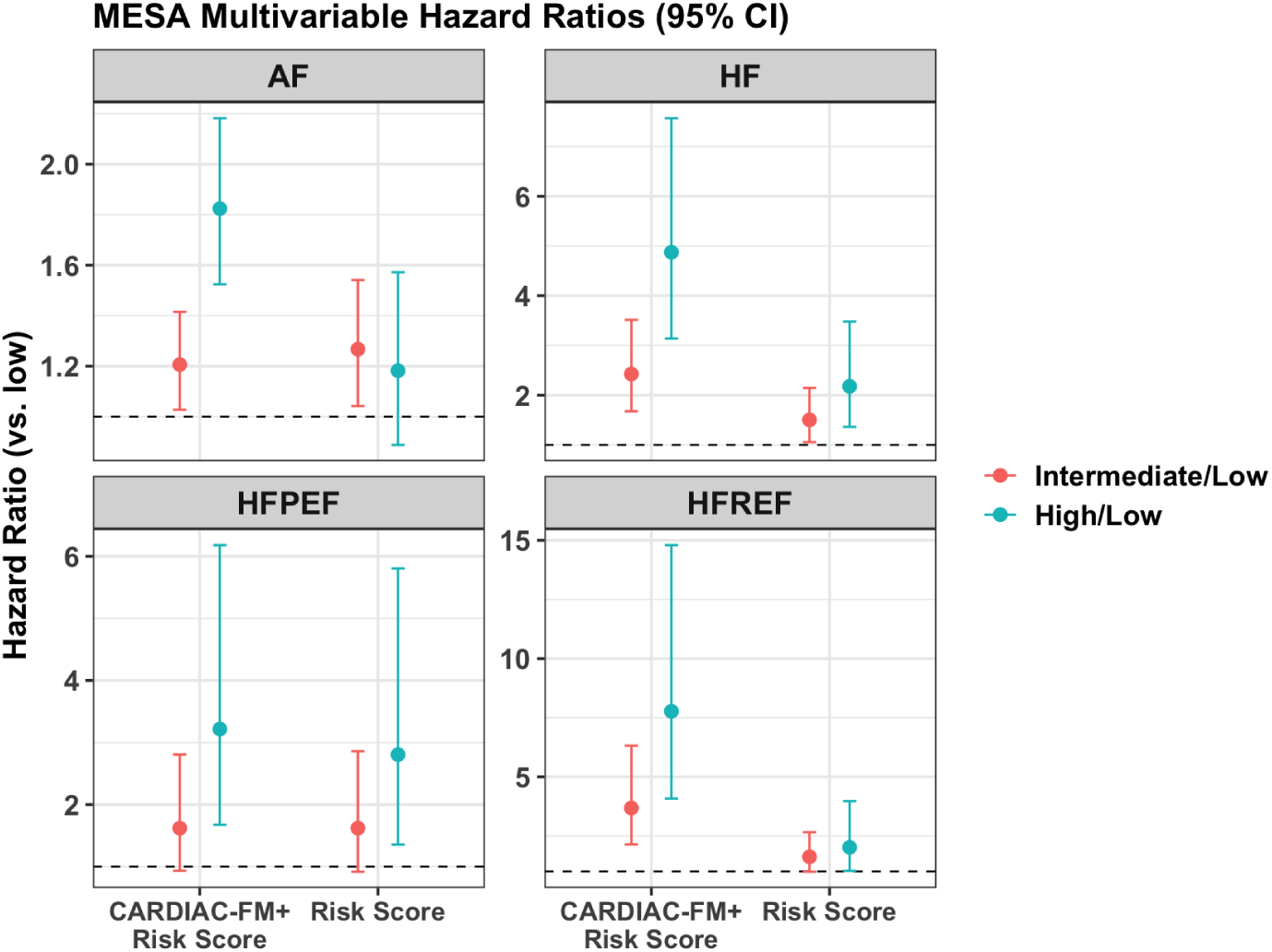
Risk stratification performance of CARDIAC-FM in external validation cohorts. Multivariable-adjusted hazard ratios for 5-year incident atrial fibrillation, heart failure, heart failure with preserved ejection fraction (HFpEF), and heart failure with reduced ejection fraction (HFrEF) in the Cardiovascular Health Study (CHS; **a**) and the Multi-Ethnic Study of Atherosclerosis (MESA; **b**). Participants were stratified into risk tertiles based on clinical risk scores alone (CHARGE-AF for atrial fibrillation; PREVENT-HF for heart failure) or zero-shot CARDIAC-FM (ECG + Risk Score). Hazard ratios were estimated using Cox proportional hazards models with the low-risk tertile as the reference and adjustment for traditional cardiovascular risk factors used in the risk score. Points indicate hazard ratios comparing the intermediate- and high-risk tertiles with the low-risk tertile; error bars indicate 95% confidence intervals obtained from the Cox proportional hazards models.

Zero-shot generalizability was also observed for time-to-event modelling based on Harrell’s C-index. CARDIAC-FM significantly outperformed all four ECG AI comparators for both atrial fibrillation and heart failure in CHS and MESA (P < 0.05; Supplementary Table 27).

We further examined whether ECG-predicted cardiac MRI phenotypes retained prognostically relevant information. In CHS, where cardiac MRI was unavailable, predicted atrial and ventricular phenotypes were associated with incident cardiovascular outcomes after adjustment for traditional risk factors (Supplementary Fig. 2 and Supplementary Table 17). In MESA, where ECG-predicted phenotypes showed moderate correlations with independently measured core-lab cardiac MRI phenotypes (r = 0.31–0.57 across seven phenotypes), we compared their associations with clinical outcomes (Supplementary Table 21). Across 28 phenotype–outcome associations, effect estimates from predicted and measured phenotypes were highly concordant (Pearson correlation of log-HRs, r = 0.93); all associations were concordant in direction, and 23 of 28 had overlapping 95% confidence intervals (Supplementary Fig. 3 and Supplementary Table 18). These findings indicate that ECG-predicted cardiac MRI phenotypes preserve clinically relevant prognostic associations observed with directly measured cardiac MRI phenotypes.

### Evaluation of multimodal pre-trained representations across a range of cardiovascular outcomes

We next evaluated whether the multimodally pretrained representation transferred to a broader range of cardiovascular outcomes. In CHS and MESA, CARDIAC-FM was fine-tuned using ECG alone for 10-year prediction of myocardial infarction, ischaemic stroke, cardiovascular death and all-cause mortality. To evaluate adaptation with limited outcome annotations, models were fine-tuned using 10% or 20% of each cohort; results for 20% fine-tuning are shown in Fig. 6, with similar findings using 10% (Supplementary Fig. 10; Supplementary Tables 11 and 12).

**Figure 6:**
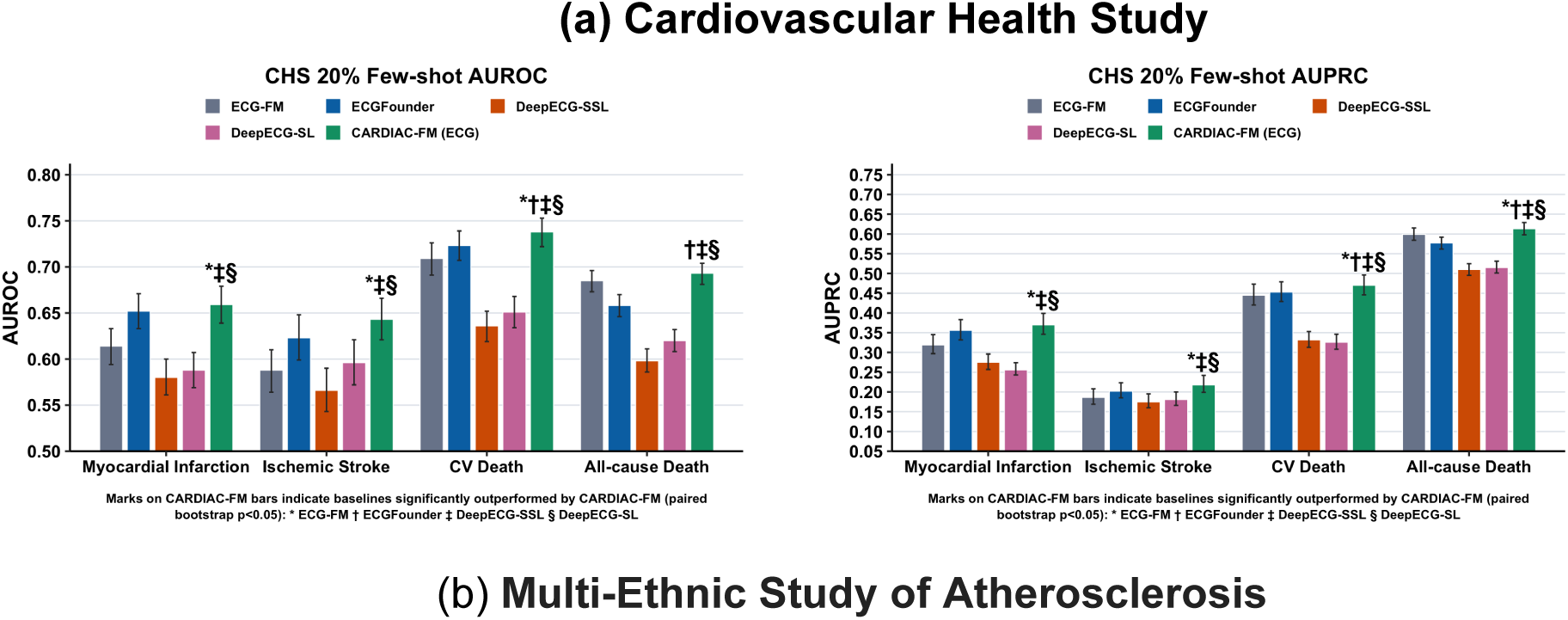
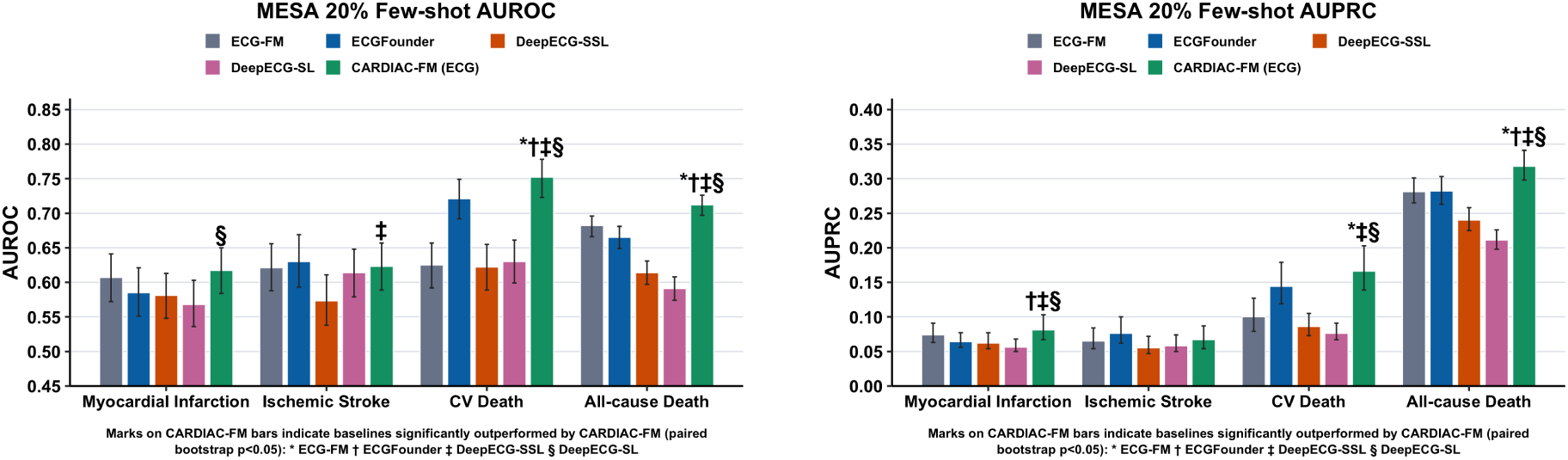
Evaluation of pre-trained representations across a broad range of cardiovascular outcomes. AUROC (left) and AUPRC (right) for CARDIAC-FM (blue) and ECG-FM (red) across four cardiovascular outcomes at a 10-year prediction horizon in CHS (**a**) and MESA (**b**). Outcomes include myocardial infarction (MI), ischaemic stroke (IS), cardiovascular death (CV death), and all-cause death (death). All models used ECG-only input and were fine-tuned using 20% of data from each cohort. CARDIAC-FM consistently outperformed ECG-FM across all outcomes in both cohorts. Error bars indicate 95% bootstrap confidence intervals. Symbols above CARDIAC-FM bars indicate statistically significant pairwise differences from comparator ECG AI models based on bootstrap (P < 0.05).

Across the four additional outcomes, CARDIAC-FM generally achieved the highest AUROC and AUPRC in both CHS and MESA, with significant improvements over multiple contemporary ECG AI comparators for most outcomes, apart from ischaemic stroke in MESA (Fig. 6). Together, these findings demonstrate that the multimodally pretrained representation can be efficiently adapted to cardiovascular outcomes beyond those used in the primary evaluation with limited task-specific annotations.

## Discussion

In this study, we developed CARDIAC-FM, a multimodal cardiovascular foundation model that combines self-supervised representation learning of ECG and cardiac MRI with cross-modal contrastive alignment. A spatiotemporal, multi-view masked autoencoder learns task-agnostic representations directly from raw cine MRI, without relying on algorithm-derived phenotype labels. Consistent with the foundation model paradigm, both encoders are pretrained without outcome labels to learn generalizable representations that can be adapted across distinct clinical tasks, including cardiovascular outcome prediction, continuous cardiac MRI phenotype prediction and censored time-to-event modelling, as well as transferred across cardiovascular outcomes and independent cohorts. A central finding is that ECG–MRI pretraining improves the ECG representation even when MRI is unavailable at deployment, while MRI provides additional predictive information when available at inference.

CARDIAC-FM generalized across populations that differed substantially from the UK Biobank imaging cohort. In CHS and MESA, the UK Biobank-trained ECG representation transferred zero-shot and consistently outperformed contemporary ECG AI models. With limited outcome fine-tuning, the representation could also be adapted to additional endpoints, including myocardial infarction, ischaemic stroke, cardiovascular mortality and all-cause mortality. Together with its performance under both fixed-horizon and time-to-event formulations, these findings suggest that multimodal pretraining learns cardiovascular features that are transferable across outcomes, prediction tasks and populations rather than being specific to a single downstream endpoint.

Importantly, CARDIAC-FM captured information complementary to established cardiovascular risk factors. Combining the ECG representation with CHARGE-AF or PREVENT-HF improved predictive performance across UK Biobank, CHS and MESA, with improvements reproduced across multiple measures of predictive performance. Risk strata defined using CARDIAC-FM remained associated with subsequent outcomes after adjustment for the clinical covariates underlying these scores, with particularly strong and consistent stratification for heart failure and HFrEF. Integrated discrimination improvement and decision-curve analyses provided complementary evidence that the learned ECG representation adds prognostic information beyond conventional clinical predictors. These findings are relevant because ECG and traditional risk factors characterize different aspects of cardiovascular health: clinical scores summarize established demographic and clinical predictors, whereas the ECG representation may capture latent electrical and structural-functional abnormalities not explicitly represented by these variables.

Beyond outcome prediction, CARDIAC-FM could be fine-tuned to predict cardiac structural and functional phenotypes from ECG. ECG-predicted cardiac MRI phenotypes showed moderate correlations with independently measured, core-lab cardiac MRI phenotypes in MESA, with the strongest correspondence for LV mass and volumes. Although these predictions are not substitutes for direct cardiac imaging, their associations with clinical outcomes closely paralleled those of measured cardiac MRI phenotypes across 28 phenotype–outcome comparisons. These findings suggest that the multimodally pretrained representation can be adapted to recover clinically relevant cardiac MRI-related information from ECG.

The framework therefore supports two complementary modes of use. In the ECG-only setting, cardiac MRI serves as privileged information during pretraining but is not required at inference, allowing information learned from a rich imaging modality to improve a comparatively inexpensive and widely available physiological signal. This configuration may be particularly relevant for population-scale applications in which cardiac MRI is impractical or unavailable. When MRI is available, the multimodal ECG+MRI configuration can additionally exploit subject-specific imaging information at inference.

Several considerations are important for interpretation. CARDIAC-FM was pretrained predominantly in UK Biobank imaging participants, and broader evaluation across healthcare systems, geographic regions and clinical populations will be needed. Although zero-shot evaluation in CHS and MESA demonstrated transfer of discrimination, absolute-risk calibration shifted across cohorts and improved after recalibration, highlighting the distinction between transportability of risk ranking and transportability of absolute risk and the potential need for local recalibration before clinical deployment. ECG-derived cardiac MRI phenotypes only moderately approximate directly measured imaging phenotypes and should not be interpreted as replacements for cardiac MRI. Furthermore, improvements in discrimination, risk stratification and decision-curve net benefit do not by themselves establish clinical effectiveness. Prospective studies will be needed to determine whether CARDIAC-FM improves clinical decisions and patient outcomes.

Overall, CARDIAC-FM demonstrates a strategy for using a deeply phenotyped imaging modality to enrich representations of a scalable physiological modality. By combining self-supervised learning within ECG and cardiac MRI with cross-modal alignment, the framework transfers across modalities, task formulations, cardiovascular outcomes and independent populations while retaining an ECG-only deployment pathway. This approach may provide a general framework for multimodal representation learning in settings where rich modalities are available during model development but cannot be assumed to be available at deployment.

## Supporting information

upplementary Figures

## Acknowledgements

UK Biobank: This research has been conducted using the UK Biobank Resource under Application Number #40713, and this work uses data provided by patients and collected by the NHS as part of their care and support. The authors would like to thank the participants and researchers involved in creating and maintaining the UK Biobank as a valuable resource for health-related research.

Cardiovascular Health Study: This Cardiovascular Health Study research was supported by NHLBI contracts HHSN268201200036C, HHSN268200800007C, HHSN268201800001C, N01HC55222, N01HC85079, N01HC85080, N01HC85081, N01HC85082, N01HC85083, N01HC85086, N01HC85084, N01HC35129, R01AG15928, R01AG20098, 75N92021D00006; and NHLBI grants U01HL080295, R01HL087652, R01HL103612, R01HL105756, R01HL120393, U01HL130114, and R01HL172803 with additional contribution from the National Institute of Neurological Disorders and Stroke (NINDS). Additional support was provided through R01AG023629 from the National Institute on Aging (NIA). A full list of principal CHS investigators and institutions can be found at CHS-NHLBI.org. The content is solely the responsibility of the authors and does not necessarily represent the official views of the National Institutes of Health.

Multi-Ethnic Study of Atherosclerosis: This research was supported by contracts 75N92020D00001, HHSN268201500003I, N01-HC-95159, 75N92020D00005, N01-HC-95160, 75N92020D00002, N01-HC-95161, 75N92020D00003, N01-HC-95162, 75N92020D00006, N01-HC-95163, 75N92020D00004, N01-HC-95164, 75N92020D00007, N01-HC-95165, N01-HC-95166, N01-HC-95167, N01-HC-95168 and N01-HC-95169 from the National Heart, Lung, and Blood Institute, grants UL1-TR-000040 and UL1-TR-001420 from NCATS, and UL1-RR-025005 from NCRR and R01-HL-127659, R01-HL-107577 and R01 HL127659 from NHLBI. The authors thank the other investigators, the staff, and the participants of the MESA study for their valuable contributions. A full list of participating MESA investigators and institutions can be found at http://www.mesa-nhlbi.org.

## Data availability

UK Biobank data are made available to researchers from research institutions as described at this site: https://www.ukbiobank.ac.uk/enable-your-research. Data for Cardiovascular Health Study and Multi-Ethnic Study of Atherosclerosis can be requested at https://internal.mesa-nhlbi.org/ and https://chs-nhlbi.org/node/6222, respectively.

## Code availability

The fully trained model and weights are available for download at https://github.com/lst627/CARDIAC-FM.

## Author contributions

T.Y., J.S.F. and S.D. designed the study. F.L., S.L., Y.Q., B.C., J.A.B., C.M.S and K.L.W. performed analyses, W.L. contributed data from MESA. B.M.P and W.L. contributed data from CHS. T.Y., F.L., S.L., B.C., and J.S.F. drafted the manuscript. All authors reviewed the manuscript and provided critical revision.

## Online Methods

### UK Biobank

The UK Biobank (UKB) is a cohort study that recruited 500,000 participants aged 40 to 69 years from the United Kingdom between 2006 and 2010^34^. The baseline assessment included comprehensive questionnaires, blood and urine collection for biochemical measurements, anthropometry, and interviews. All participants provided electronic informed consent at the UK Biobank assessment centers. The UKB received ethics approval from the North West Multi-Centre Research Ethics Committee.

Longitudinal follow-up for health outcomes of atrial fibrillation, and heart failure was performed through linkages with national databases on hospitalizations, primary care records, and death registries. Incident atrial fibrillation events were identified from self-reports, nurse interviews, hospital admission codes, death register cause-of-death codes, and UK Biobank first occurrence data. Heart failure was identified solely from diagnosis codes and cause-of-death codes (Supplementary Table 22).

We defined binary prediction outcomes at a 5-year time horizon following the imaging visit. For each participant, we coded atrial fibrillation status as positive if the participant had no prior diagnosis of atrial fibrillation at the imaging visit and developed incident atrial fibrillation within 5 years, as negative if the participant completed 5-year follow-up without an event, and as missing otherwise. We excluded participants with prevalent atrial fibrillation at the imaging visit from atrial fibrillation prediction analyses. We defined heart failure analogously and excluded participants with prevalent disease at the imaging visit from respective analyses.

Participants whose imaging visits occurred after the UKB data censoring dates were excluded. Follow-up for this study extended through 2022. Hospital data were available until October 2022 for England, August 2022 for Scotland, and May 2022 for Wales. Death record data was censored in November 2022 across all sites.

Traditional cardiovascular risk factors were measured at the imaging visit or, when unavailable, at the most recent prior visit. Covariates included age, sex, height, weight, body mass index (BMI), systolic and diastolic blood pressure, total cholesterol, high-density lipoprotein cholesterol, estimated glomerular filtration rate (eGFR; calculated using the 2009 Chronic Kidney Disease Epidemiology Collaboration creatinine equation), diabetes mellitus, current smoking status, prevalent myocardial infarction, and current use of antihypertensive and lipid-lowering medications. These variables comprise the CHARGE-AF and American Heart Association PREVENT-HF (Predicting Risk of Cardiovascular Disease EVENTs equation for heart failure).

### Cardiovascular Health Study

The Cardiovascular Health Study (CHS) is a prospective cohort study of risk factors for cardiovascular diseases in adults aged 65 years and older, recruited from four field centers in the United States (Sacramento, CA; Hagerstown, MD; Winston-Salem, NC; Pittsburgh, PA)^35^. Between June 1989 and June 1990, 5,201 participants were recruited from random samples of Medicare eligibility lists. An additional 687 Black participants were recruited between November 1992 and June 1993. Study participants were seen in the clinic annually between enrollment and 1998-99 and were contacted by telephone at 6-month intervals through June 2015 to collect information about health status, hospitalizations, and medication use^36^. The study was approved by the institutional review boards of all participating sites, and all study participants provided informed consent.

Atrial fibrillation events were identified using a combination of ECG findings from clinic visits through 1999, hospital discharge records indicating atrial fibrillation, and Medicare inpatient, outpatient, and physician (carrier) claims data. Resting 12-lead ECGs were performed at baseline and follow-up visits, with centralized readings to classify atrial fibrillation according to standardized criteria. Resting 12-lead ECGs from baseline and Year 7 were included in our analyses. For atrial fibrillation identified through Medicare data, diagnosis required either ECG evidence, one inpatient claim, or two outpatient or physician claims within 365 days using International Classification of Diseases, Ninth Revision (ICD-9) codes 427.31 or 427.32 in any diagnostic position.

Assessment of heart failure events in CHS has been previously described^37^. All potential heart failure events were identified through in-person visits or telephone interviews. These events were then adjudicated by the central events committee. Adjudication of heart failure was based on a physician diagnosis of heart failure, documented symptoms and signs of heart failure, use of medical treatment for heart failure, and supportive findings on diagnostic testing.

Methods for clinical ischemic stroke adjudication in CHS have been previously described^38^. In summary, an expert events committee, including neurologists from the study sites and CHS coordinating center, along with a neuroradiologist from the imaging reading center, reviewed the cases. Ischemic stroke was characterized by the sudden onset of a focal neurological deficit persisting for at least 24 hours, supported by compatible imaging findings on computed tomography scan or cardiac magnetic resonance imaging, and without evidence of intracranial hemorrhage or a nonvascular underlying cause.

Myocardial infarction (MI) was adjudicated to have occurred in the presence of evolving Q-wave MI or cardiac pain plus abnormal enzymes and either an evolving ST-T pattern or new left bundle branch block.

Details of surveillance methods and disease classification in CHS have been described previously^39^. Since participants were enrolled in the study, deaths were identified through phone calls, annual clinic visits, or local obituaries. Cause of death was adjudicated by a centralized adjudication committee, which reviewed information from obituaries, death certificates, medical records, and proxy interviews^27^. Cardiovascular death is defined as death from coronary artery disease.

### Multi-Ethnic Study of Atherosclerosis

The Multi-Ethnic Study of Atherosclerosis (MESA) is a prospective, population-based observational cohort study of 6,814 men and women representing four racial/ethnic groups (white, African American, Hispanic, and Chinese-American), aged 45–84 years and free of clinical cardiovascular disease at enrollment^40^. Study participants were recruited in 2000-2 at 6 field centers in the US (Baltimore, MD; Chicago, IL; Forsyth County, NC; Los Angeles, CA; New York, NY; and St. Paul, MN). At telephone contacts every 9–12 months during follow-up, participants were asked to identify new hospitalizations and diagnoses, and medical records were obtained. Institutional review boards of all field centers approved the study protocol, and all participants gave written informed consent. Resting 12-lead ECGs from Exams 1 and 5 were included in our analyses.

Clinically recognized atrial fibrillation was identified by an ICD hospital discharge diagnosis code (version 9: 427.31 or 427.32; version 10: I48) in any position; and for those enrolled in fee-for-service Medicare, by an inpatient, outpatient, or physician claim with an atrial fibrillation code^41^. A panel reviewed medical records and adjudicated heart failure events according to standardized criteria. The present study included probable or definite heart failure events. Probable heart failure required heart failure symptoms, a physician diagnosis of heart failure, and treatment. Definite heart failure required at least one objective feature of heart failure, including abnormalities on chest X-ray, echocardiography, or ventriculography.

Methods for clinical ischemic stroke adjudication in MESA has been previously described^42^. In brief, information about all new cardiovascular conditions, hospital admissions, cardiovascular outpatient diagnoses, treatments, and deaths were obtained. Two vascular neurologists from the MESA study events committee independently reviewed all medical records for end point classification and assignment of incidence dates. They reviewed and classified stroke as present if there was a focal neurologic deficit lasting 24 hours or until death or, if <24h, there was a clinically relevant lesion on brain imaging and there was no nonvascular cause. Patients with focal neurological deficits secondary to brain trauma, tumor, infection, or other nonvascular cause were excluded. Ischemic strokes were distinguished from hemorrhagic stroke using findings on imaging, surgery, autopsy, or some combination of these. Ischemic stroke subtypes were assigned based on an extension of the Trial of Org 10172 in Acute Stroke Treatment (TOAST) criteria to try to reduce the number classified as undetermined.

A myocardial infarction in MESA required either abnormal cardiac biomarkers (2 times upper limits of normal) regardless of pain or ECG findings; evolving Q waves regardless of pain or biomarker findings; or a combination of chest pain, ST-T evolution or new left bundle branch block, and biomarker levels 1 to 2 times upper limits of normal^43^.

Hard cardiovascular disease events in MESA were required to be symptomatic and included myocardial infarction (MI), resuscitated cardiac arrest, stroke, and cardiovascular death. Incident heart failure events were recorded. All deaths were identified. For potential cardiovascular deaths, cause was assigned through committee review. For other deaths, the underlying cause was obtained through state or city vital statistics departments^44^.

### Cardiac magnetic resonance imaging

The UK Biobank Imaging Study, initiated in 2014, includes magnetic resonance imaging of the brain, abdomen, and heart^45^. The first 5,065 cardiac MRI scans were manually analyzed, and LA contours at end-systole and end-diastole were traced^46^. Bai et al^47^. used these annotated images to develop a fully automated convolutional network pipeline for large-scale quantification of cardiac structure and function. The algorithm’s accuracy was evaluated using technical metrics (Dice coefficient, contour distance, and Hausdorff distance) and validated by clinical experts. This publicly available algorithm was applied to derive MRI parameters, including left ventricular ejection fraction (LVEF), end-diastolic and end-systolic volumes (LVEDV, LVESV), left ventricular mass (LVM), left atrial emptying fraction (LAEF), and minimum and maximum left atrial volumes (LAVmin, LAVmax).

In MESA, cardiac MRI phenotypes were independently measured rather than derived using the automated pipeline applied in UK Biobank. Cine cardiac MRI was analyzed using established MESA protocols. For left ventricular measurements, trained MESA Core Lab readers delineated endocardial and epicardial borders, with contours reviewed and corrected by cardiac MR physicians; left atrial volumes were derived from cine cardiac MRI using validated contour-based methods. Left atrial and left ventricular volumes, mass and ejection fraction obtained from these measurements were used as independent reference values against which ECG-predicted cardiac MRI phenotypes were evaluated in MESA^48^.

**Cardiac MRI preprocessing.** Our cardiac MRI preprocessing follows the pipeline proposed by Wang et al.^20^, extended to a third view. For each participant we processed three cine sequences: the two-chamber (2CH) and four-chamber (4CH) long-axis views and the short-axis (SAX) stack, together with their automated chamber segmentations. As in Wang et al., the SAX view was represented by three slices: the mid-ventricular slice and the slices two positions above and below it, while the 2CH and 4CH views each contributed a single slice. Each volume was resampled to a common in-plane resolution of 0.994 × 0.994 mm, which is done by linear interpolation for images, and nearest-neighbour for segmentation label maps. We then localized the heart from the segmentation as the bounding box enclosing the relevant chambers: the left atrium for the two-chamber view, the left atrium and left ventricle for the four-chamber view, and the left-ventricular cavity, myocardium and right ventricle for the short-axis view. This box was taken as the union across the selected slices and all cardiac phases, then expanded by a 10% margin. We cropped each frame to this box, zero-padded it to preserve the aspect ratio, and resized it to 224 × 224 pixels. Cine sequences were temporally subsampled with a stride of two, reducing 50 frames to 25. Following Wang et al., we clipped pixel intensities at the 0.1st and 99.9th percentiles, rescaled them linearly to the range 1 to 255, and standardized them to zero mean and unit variance within each volume. This produced sequences of 25 frames at 224 × 224 pixels for each participant, with a single slice for each long-axis view and three slices for the short-axis stack.

### Self-supervised cardiac MRI encoder

We developed a multi-view convolution–transformer masked autoencoder in the family of masked autoencoders^49^, and inspired by the multi-modal masked autoencoder MultiMAE^50^ and cardiac masked autoencoder CineMA^51^, in order to handle the multi-view nature of MRI data. In contrast to CineMA, which samples a single cine frame per study and performs two-dimensional image encoding, our model preserves the temporal dimension and performs spatiotemporal masked autoencoding using VideoMAE-style tube masking^52^. This design enables the encoder to learn cardiac motion in addition to cardiac anatomy, which is particularly relevant for subsequent alignment with the temporally resolved ECG signal.

After preprocessing, for each participant we have three views of cardiac MRI, which are two-chamber (2CH) and four-chamber (4CH) long-axis, and three mid-ventricular short-axis (SAX) slices, each forming a (1, T, H, W) volume with T=25 frames and H=W=224. At each training step a continuous 8-frame window is sampled uniformly at random from the 25-frame cine as a temporal augmentation, and the center 8 frames are used at inference.

Each view is tokenized by a ConvMAE-style convolutional stem^53^ with view-specific weights that downsamples each frame spatially by 16× and pools every two adjacent frames into one temporal token, yielding a 4×14×14 grid (784 tokens) per single-slice stream (2CH, 4CH, and each of the three SAX slices, i.e. five streams in total). Fixed 3-D sin–cos positional embeddings, learned view-type embeddings (2CH/4CH/SAX), and learned SAX-slice embeddings are added.

During self-supervised pretraining, we mask 75% of tokens using tube masking: a random 25% of the 14×14 spatial locations is retained and the pattern is shared across the temporal axis, so every temporal token at a retained location is visible and every temporal token at a masked location is hidden. This strategy of masking all temporal tokens at the same location prevents reconstruction by copying from an adjacent visible frame at the same location and forces the model to use spatial context and cross-view information.

Visible tokens from all five streams are concatenated and pass through a ViT-base encoder (width 768, 12 layers, 12 heads), integrating information across views, slices, and time. A lightweight decoder (width 512, 8 layers, 16 heads) with per-view linear pixel heads reconstructs the masked patches, minimizing the normalized-pixel mean-squared error over masked patches, averaged across views.

At inference the cardiac MRI encoder runs unmasked, and we read out a per-view pooled representation: the fused tokens are mean-pooled within each view and concatenated into a 2,304-dimensional (3 × 768) study-level embedding. This embedding is linearly projected and aligned with the ECG-FM embedding via the InfoNCE loss in the subsequent multimodal learning step.

### Electrocardiograms

In UK Biobank, each ECG trace comprises 5,000 uniformly sampled data points per lead over 10 seconds at 500 Hz. We parsed 12-lead ECG signals, including all standard leads (I, II, III, aVR, aVL, aVF, V1–V6).

We excluded ECGs with poor quality, suspected lead reversal, undetermined rhythm, or implausible ventricular rates (<20 or >140 bpm). We also excluded ECGs showing atrial fibrillation, atrial flutter, or pacemaker activity. After quality control, 67,661 samples with valid 12-lead ECG data were retained for analysis.

To remove baseline wander, we applied locally weighted scatterplot smoothing (LOWESS) independently to each lead. For each 5,000-point trace, we fitted a LOWESS curve using the signal as the response variable and sample index as the predictor with a smoothing fraction of 0.1. The resulting smoothed curve, representing low-frequency drift, was subtracted from the raw signal to obtain baseline-corrected ECG data.

In CHS and MESA, resting 10-second 12-lead ECGs were recorded at 500 Hz using MAC PC ECG machines (Marquette Electronics, Milwaukee, WI) at baseline and selected follow-up visits. All ECGs were centrally reviewed for technical errors and quality at the Epidemiological Cardiology Research Center (EPICARE), Wake Forest University School of Medicine (Winston-Salem, NC), using General Electric 12-SL software (GE Healthcare, Milwaukee, WI) on MUSE and Magellan workstations. In the ECG signals from both the CHS and MESA cohorts, no significant baseline wander was observed. ECG signals from both cohorts showed no significant baseline wander. CHS ECG amplitudes were rescaled to align with UK Biobank reference values, whereas MESA ECG amplitudes were already comparable and required no adjustment.

### Multimodal AI development

**CARDIAC-FM framework.** The CARDIAC-FM framework comprised a two-stage pipeline. In the first stage, modality-specific encoders extracted features from ECG and MRI inputs, which were aligned in a shared latent space using symmetric InfoNCE contrastive loss^30^. The ECG encoder was initialized from the pretrained ECG Foundation Model (ECG-FM), whereas the cardiac MRI encoder was the self-supervised spatiotemporal multi-view masked autoencoder described above. Importantly, cardiac MRI encoder pretraining used only cine image reconstruction and did not use algorithm-derived cardiac MRI phenotypes, clinical diagnoses or subsequent clinical outcomes as supervision.

The ECG-FM encoder comprises a multi-layer convolutional feature extractor followed by a BERT-Large transformer encoder (https://github.com/bowang-lab/ECG-FM). The cardiac MRI encoder jointly represents information across long-axis and short-axis views and across cardiac phases using the convolution–transformer architecture described above. Linear projection layers mapped the ECG and cardiac MRI representations to a common embedding dimension for cross-modal alignment. During contrastive multimodal training, both the ECG and cardiac MRI encoders were fully fine-tuned.

In the second stage, a prediction layer was added to the learned representations and the entire model was fine-tuned with supervised learning for clinical outcome prediction. The final model accepts two input configurations—ECG-only or combined ECG+MRI—enabling broad clinical applicability across settings with varying data availability.

**Loss objectives for multimodal pretraining.** The learned representations were trained using symmetric InfoNCE loss across modalities^30^. Consider a batch containing *B* paired samples, where each sample consists of one ECG input and one MRI input, denoted by {(*x_i_^ECG^*, *x_i_^MRI^*)}*_i_*_=1_*^B^*. The modality-specific encoders produce feature representations:

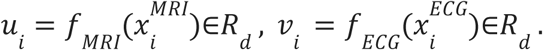

The similarity function between representations is defined as:

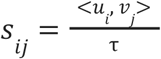

Where τ denotes the temperature parameter. The InfoNCE Loss for contrastive learning is then computed as:

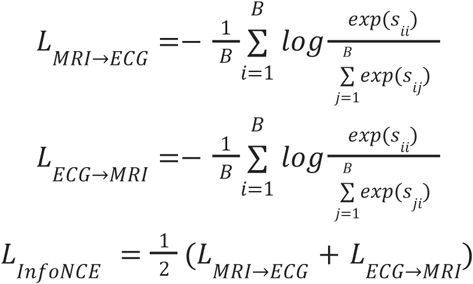

This loss is computed across ECG-MRI pairs within each batch, pulling together embeddings from paired modalities of the same subject while pushing apart embeddings from different individuals. This contrastive learning step established the foundation for subsequent fine-tuning on downstream prediction tasks.

**Training Details.** The MRI encoder pretraining was performed on four NVIDIA H200 GPUs with a batch size of 128 (16 per GPU × 2 gradient accumulation × 4 GPUs) for 200 epochs with early stopping at patience 7 using the AdamW optimizer (β₁ = 0.9, β₂ = 0.95, ε = 1×10⁻⁶, weight decay = 0.05). Learning rate followed a cosine annealing schedule with base rate 1.5×10⁻⁴ and warm-up length of 10 epochs. The mask ratio is set to be 0.75.

Multimodal pretraining was performed on four NVIDIA H200 GPUs with a batch size of 32 (8 per GPU × 4 GPUs) for 50 epochs using the AdamW optimizer (β₁ = 0.9, β₂ = 0.98, ε = 1×10⁻⁶, weight decay = 0.05). Learning rate followed a cosine annealing schedule with base rate 1×10⁻⁵ and warm-up length of 50 steps. The temperature parameter in the InfoNCE loss was trainable and initialized at 0.07.

For downstream binary outcome prediction, prediction heads were appended to the pretrained representations and the models were fine-tuned on UK Biobank. All encoder parameters were trainable during fine-tuning. Training uses batch size of 32 for 20 epochs with early stopping at patience 3 using the AdamW optimizer (β₁ = 0.9, β₂ = 0.98, ε = 1×10⁻⁶, weight decay = 0.01) with a linear warm-up schedule. Base learning rate is 5×10⁻⁶ for ECG-only model and 1×10⁻⁶ for the ECG+MRI model, with warm-up length 50 steps. Model checkpoints were selected based on the highest AUROC on the validation set. No hyperparameter tuning was performed beyond optimizer and learning rate scheduler selection.

To further evaluate the representation on a task with a different objective and metric, we additionally modeled time to atrial fibrillation and heart failure as a time-to-event outcome that is subject to right censoring. Participants with prevalent disease were excluded for the corresponding outcome. A single linear head on the ECG encoder produced a continuous log-hazard, and the encoder was fine-tuned by minimizing the Cox proportional-hazards partial likelihood^54^. Models were selected on Harrell’s C-index and evaluated by the test-set C-index on the UK Biobank test set. The survival models trained by UK Biobank data were then applied zero-shot to CHS and MESA, and evaluated by the C-index against each cohort’s time-to-event outcomes with prevalent cases excluded.

For the risk factors in the clinical risk models, all predictors except sex contained varying degrees of missingness (approximately 1–18%). We applied multiple imputation using chained equations (MICE) before model fitting, generating four imputed datasets. The CHARGE-AF and PREVENT-HF scores were then evaluated in each of the four completed datasets and the four scores are averaged to give a single risk score for each participant.

We additionally evaluated clinical models in which these coefficients were re-estimated using UK Biobank. Logistic regression models containing the individual CHARGE-AF variables for atrial fibrillation or PREVENT-HF variables for heart failure were fitted using the UK Biobank training set. These fitted models were evaluated in the held-out UK Biobank test set and subsequently transferred without refitting to CHS and MESA, mirroring the external-validation procedure used for CARDIAC-FM. The result is in the Supplementary Table 26.

**Data used for pretraining and fine-tuning.** For self-supervised pretraining, we used eligible ECG and cardiac MRI data from both the baseline and repeat imaging visits in UK Biobank. When multiple visits were available for the same participant, all visits from that participant were assigned to the same data partition to prevent information leakage. For downstream outcome-specific fine-tuning, only the baseline imaging visit was used. Repeat imaging visits were excluded from fine-tuning because they did not have sufficient subsequent follow-up for the prespecified prediction horizons.

**Additional Baseline ECG models.** Except for ECG-FM, we compared CARDIAC-FM against three other independently developed ECG models. These baselines were: ECGFounder, a large-scale supervised ECG foundation model based on a one-dimensional convolutional network (Net1D)^55^; DeepECG-SL, a supervised EfficientNet-V2 model trained on 77 diagnostic labels^56^; and DeepECG-SSL, a self-supervised wav2vec2 model with contrastive multi-segment coding^57^. For binary outcome prediction every model was fully fine-tuned on the same UK Biobank training data with the same training configuration as finetuning of CARDIAC-FM: AdamW optimizer, learning rate 5×10⁻⁶, early-stopping criterion (validation AUROC with patience 3), and 20 epochs. For the survival task the same models were fine-tuned with the DeepSurv^58^ objective. Baselines were applied zero-shot and few-shot to CHS and MESA using the same protocols as CARDIAC-FM.

**Subgroup analysis.** We performed subgroup analyses stratified by sex and age. In UK Biobank and MESA, age subgroups were defined as <65 versus ≥65 years. Because CHS enrolled an older population, with participants aged 65 years or older at enrollment, we instead used <75 versus ≥75 years to provide a more informative age stratification within that cohort. Model performance was evaluated separately within each subgroup.

### Statistical Analysis

**Confidence Intervals.** For every metric we report the 95% confidence interval obtained by nonparametric bootstrap resampling of test participants. For the binary discrimination metrics (AUROC, AUPRC) we used a stratified bootstrap in which cases and controls were resampled separately to their observed counts, so that each resample preserves the event rate of the test set, which avoids the unstable metric values that are due to the rareness of some outcomes. For the time-to-event analyses, participants were resampled without stratification. Interval bounds are the 2.5th and 97.5th percentiles of the resulting bootstrap distribution. For the external validation cohort (CHS and MESA), intervals were computed within CHS and MESA separately, applying the identical procedure to each cohort’s test population.

**Bootstrap p-values.** Models were compared to one another with a paired bootstrap. Within each bootstrap resample, both models were evaluated on the same resampled participants. For each resample we recorded the difference in performance between the two models, using whichever metric the comparison concerned. Repeating this across all resamples yields a distribution of performance differences, which is the sampling distribution of the model-versus-model gap. A two-sided p-value was then obtained from the position of zero within that distribution: we took the proportion of resamples in which the difference favoured the first model or was exactly zero, and the proportion in which it favoured the second model or was exactly zero, and doubled the smaller of the two.

**Cox proportional hazards analysis across risk strata.** We performed Cox regression analyses to evaluate the prognostic value of three model configurations: risk scores alone, CARDIAC-FM (ECG + risk scores), and CARDIAC-FM (ECG + MRI + risk scores). For each model and outcome (atrial fibrillation and heart failure), predicted risk scores (averaged across four random seeds) were stratified into tertiles representing high-, intermediate-, and low-risk groups, with the low-risk tertile serving as the reference. We estimated hazard ratios and 95% confidence intervals for the intermediate- and high-risk groups using univariable and multivariable Cox proportional hazards models. Multivariable models adjusted for the traditional risk factors included in the corresponding established clinical risk score. Multivariable-adjusted estimates were reported as the primary results. Because these models included multiply imputed risk factors, we calculated confidence intervals using Rubin’s rules.

**Cox proportional hazards analysis across MRI features.** We performed Cox regression analyses to evaluate the prognostic value of cardiac MRI features, including left atrial volumes (LAVmax, LAVmin), left atrial emptying fraction (LAEF), left ventricular volumes (LVEDV, LVESV), left ventricular ejection fraction (LVEF), and left ventricular mass (LVM). We estimated hazard ratios using Cox proportional hazards models adjusted for traditional risk factors. Because these models included multiply imputed risk factors, we calculated confidence intervals using Rubin’s rules.

In UK Biobank, Cox models for atrial fibrillation and heart failure were constructed separately using two distinct sets of predictors: measured MRI features and CARDIAC-FM-predicted MRI features derived from ECG. Hazard ratios were estimated from both model sets to compare prognostic performance (Supplementary Table 19). In the Cardiovascular Health Study and Multi-Ethnic Study of Atherosclerosis, Cox models were fitted for atrial fibrillation, heart failure and their respective subtypes using CARDIAC-FM-predicted MRI features derived from models trained on UK Biobank. In MESA, where cardiac MRI was performed, models using measured MRI features were also constructed, enabling direct comparison between predicted and measured structural parameters.

**Calibration analysis.** Calibration of the predicted 5-year probabilities was assessed on the UKBB test set and the external cohorts (CHS and MESA). Because the raw output of a fine-tuned neural network is not a calibrated probability, each model’s scores were corrected by logistic recalibration: fitting a logistic regression using the raw score on the evaluation cohort before evaluating with any probability-dependent metric (IDI, decision-curve net benefit). How well the raw models are calibrated was summarized by the calibration intercept and slope, and the Brier score, and displayed with reliability diagrams, which shows the mean predicted probability versus observed event frequency in deciles^59^.

**Reclassification analysis.** Our primary reclassification analysis focused on the integrated discrimination improvement (IDI). Following Pencina et al.^60^, IDI was calculated as the change in discrimination slope for the new model relative to the base model, where the discrimination slope is the mean predicted risk among participants who developed the outcome minus the mean predicted risk among participants who did not. In the primary analysis, the comparison was between the CARDIAC-FM plus risk score versus the risk score alone, so the IDI estimates the incremental improvement from adding the ECG representation to the clinical risk score. We also computed pairwise IDI contrasts between CARDIAC-FM and alternative ECG models. These model-to-model IDI contrasts quantify differences in calibrated risk separation, serving as an alternative metric for differences in AUROC and AUPRC.

**Decision-curve analysis.** Decision-curve analysis^58^ evaluates whether using a prediction model to classify participants as high risk would provide more clinical benefit than default strategies such as flagging everyone or flagging no one. Across a range of risk thresholds, participants with predicted risk above the threshold were considered high risk, and each strategy was scored by rewarding true event detection while penalizing unnecessary high-risk classification. The size of this penalty depends on the chosen threshold, reflecting the clinical trade-off between missing future events and acting on participants who would not develop the outcome. We compared net benefit curves for the risk score alone, CARDIAC-FM plus risk score, and two reference strategies: treat-all (predicting all individuals as positive) and treat-none (predicting all individuals as negative).

