## Supplementary material for "CARDIAC-FM: A Generalizable Multimodal Foundation Model Integrating ECG and Cardiac MRI": upplementary Figures

### Supplementary Figures for “CARDIAC-FM: A Generalizable Multimodal Foundation Model Integrating ECG and Cardiac MRI”

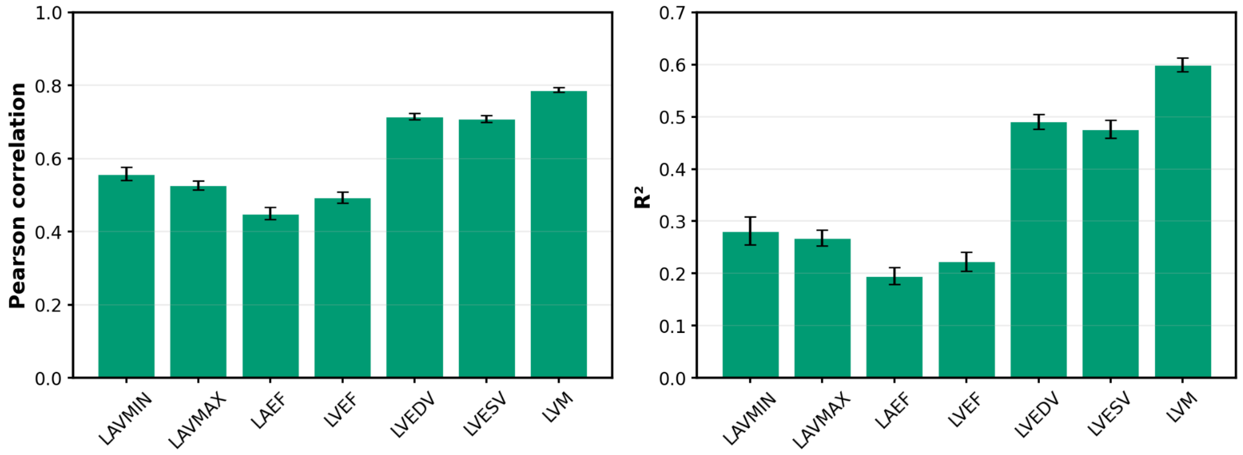


**Supplementary Figure 1. Prediction of MRI-derived cardiac phenotypes from ECG using CARDIAC-FM in UK Biobank.** Pearson correlation coefficients (left) and coefficients of determination ($R^2$; right) for ECG-based prediction of left atrial minimum volume (LAVmin), left atrial maximum volume (LAVmax), left atrial emptying fraction (LAEF), left ventricular ejection fraction (LVEF), left ventricular end-diastolic volume (LVEDV), left ventricular end-systolic volume (LVESV), and left ventricular mass (LVM). Error bars represent 95% bootstrap confidence intervals.

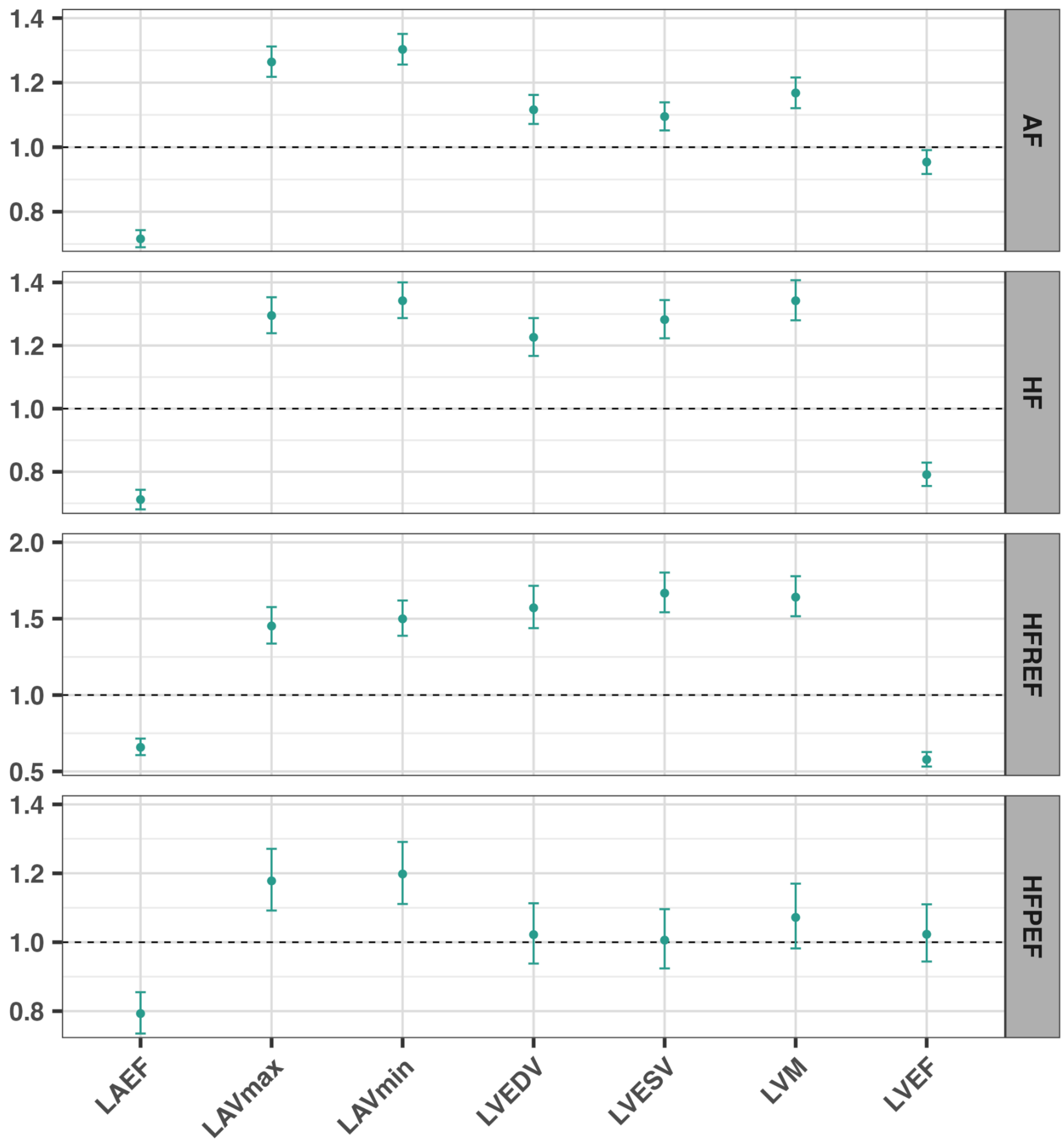


**Supplementary Figure 2. Hazard ratios (95% CI) for zero-shot CARDIAC-FM-predicted cardiac MRI features in relation to incident cardiovascular events in the CHS cohort.** Predicted cardiac MRI parameters include LA ejection fraction (EF), maximum volume, and minimum volume, as well as LV end-diastolic volume (EDV), end-systolic volume (ESV), mass, and EF. Hazard ratios were estimated using Cox proportional hazards models adjusted for traditional cardiovascular risk factors and are expressed per unit increase in each predicted cardiac MRI parameter. The dashed line indicates the null value (HR = 1). Error bars represent 95% confidence intervals.

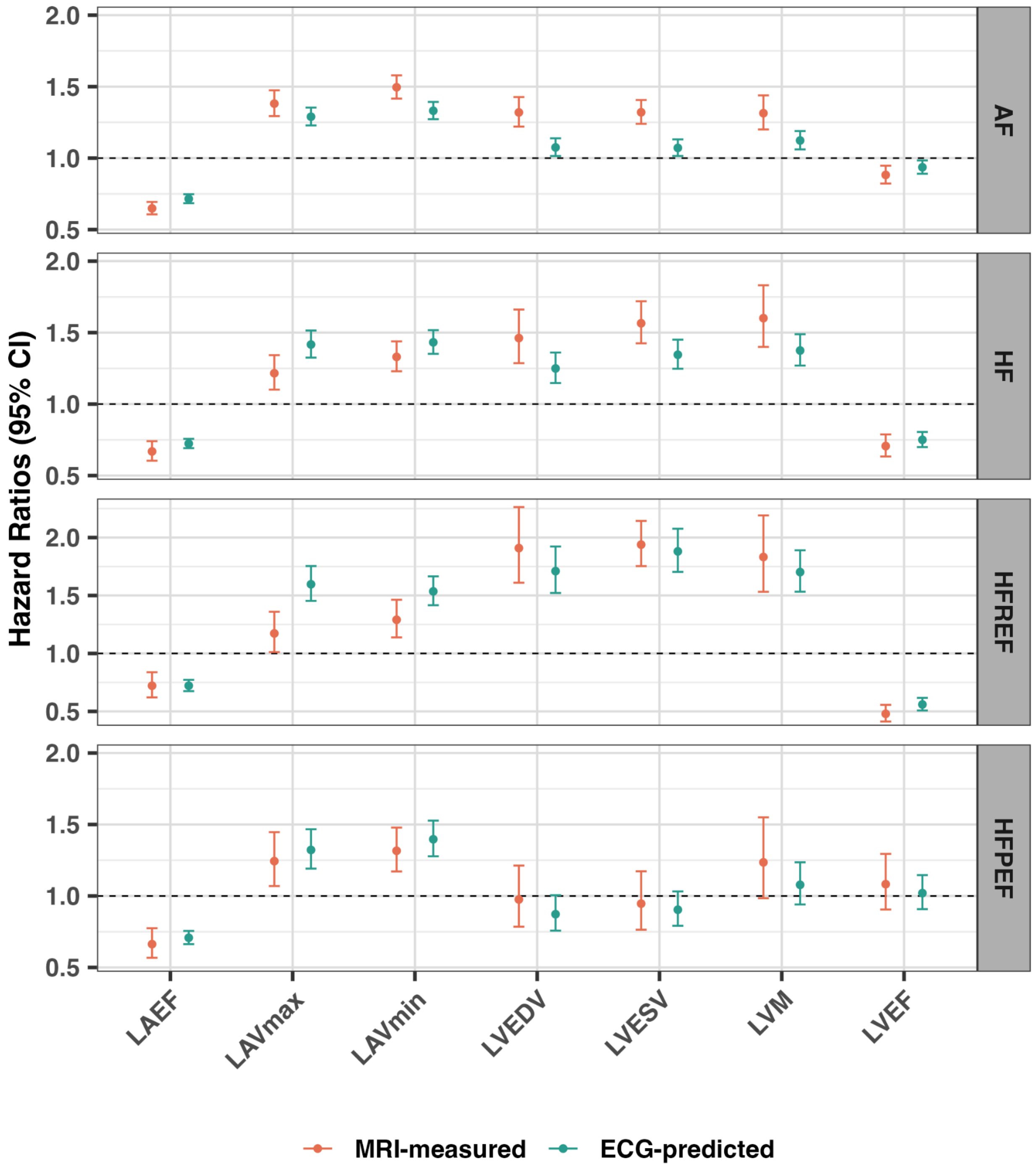


**Supplementary Figure 3. Hazard ratios (95% CI) for zero-shot CARDIAC-FM-predicted and MRI-measured cardiac MRI features in relation to incident cardiovascular events in the MESA cohort.** cardiac MRI parameters include LA ejection fraction (EF), maximum volume, and minimum volume, as well as LV end-diastolic volume (EDV), end-systolic volume (ESV), mass, and EF. Hazard ratios were estimated using Cox proportional hazards models adjusted for traditional cardiovascular risk factors and are expressed per unit increase in each predicted cardiac MRI parameter. The dashed line indicates the null value (HR = 1). Error bars represent 95% confidence intervals.

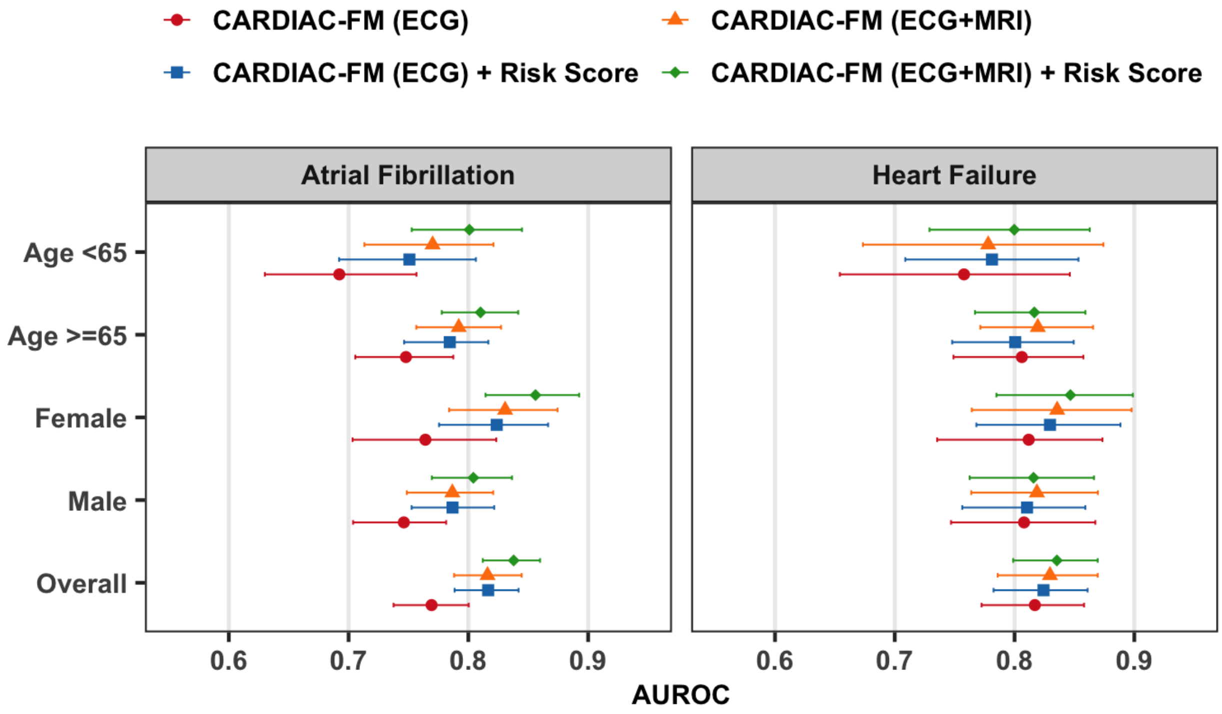


**Supplementary Figure 4. Subgroup analysis of CARDIAC-FM performance in the UK Biobank.** AUROC for 5-year prediction of incident atrial fibrillation (left) and heart failure (right), stratified by age (<65 versus ≥65 years) and sex. Error bars indicate 95% bootstrap confidence intervals.

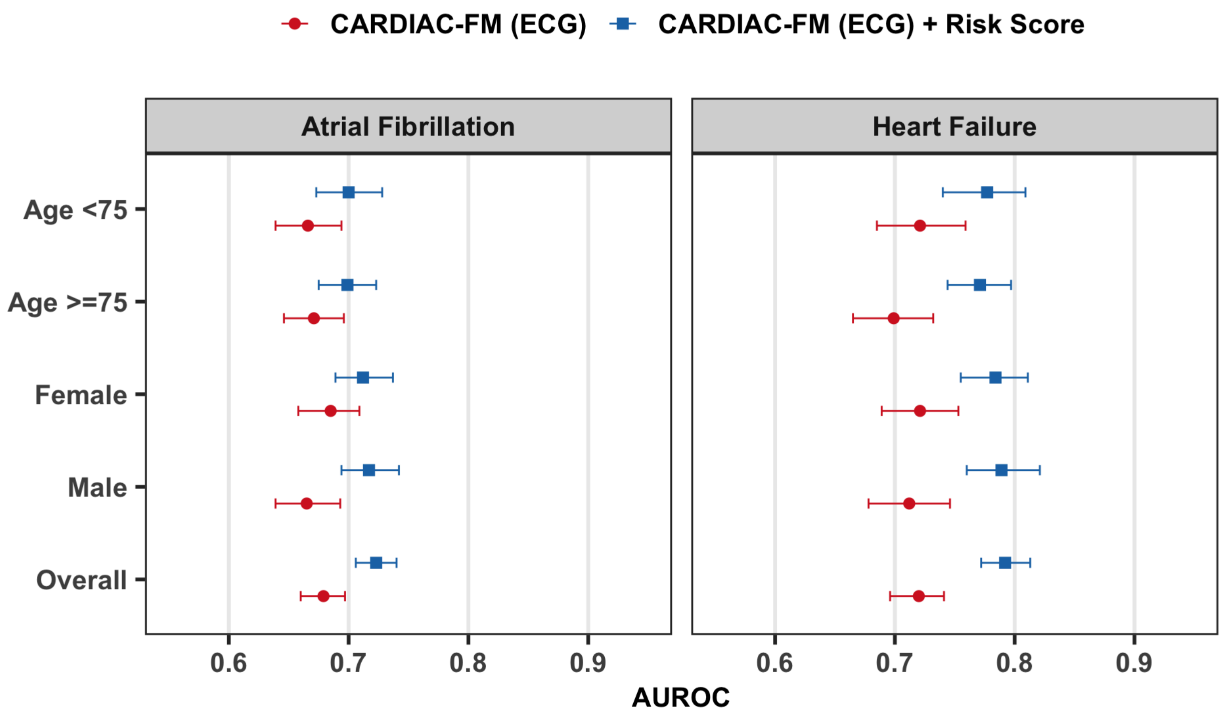


**Supplementary Figure 5. Subgroup analysis of CARDIAC-FM performance in CHS.** AUROC for 5-year prediction of incident atrial fibrillation (left) and heart failure (right), stratified by age (<75 versus ≥75 years) and sex. Red points indicate CARDIAC-FM with ECG; blue points indicate CARDIAC-FM with ECG and clinical risk score. Error bars indicate 95% bootstrap confidence intervals.

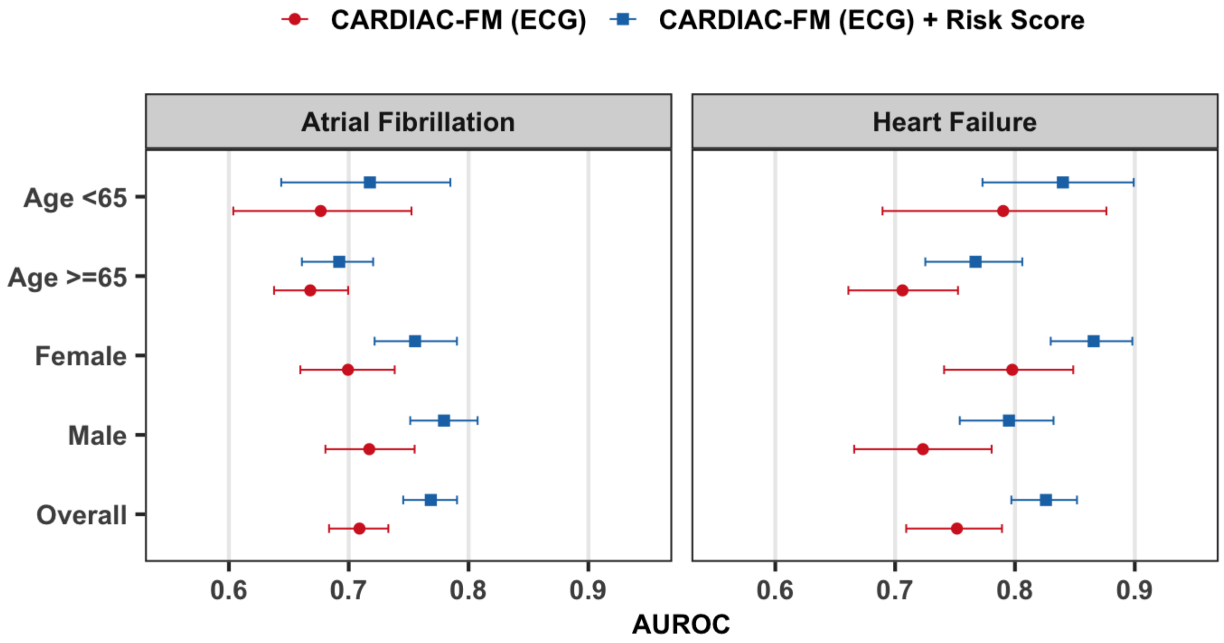


**Supplementary Figure 6. Subgroup analysis of CARDIAC-FM performance in MESA.** AUROC for 5-year prediction of incident atrial fibrillation (left) and heart failure (right), stratified by age (<65 versus ≥65 years) and sex. Red points indicate CARDIAC-FM with ECG; blue points indicate CARDIAC-FM with ECG and clinical risk score. Error bars indicate 95% bootstrap confidence intervals.

***Calibration Plots and Decision Curves***

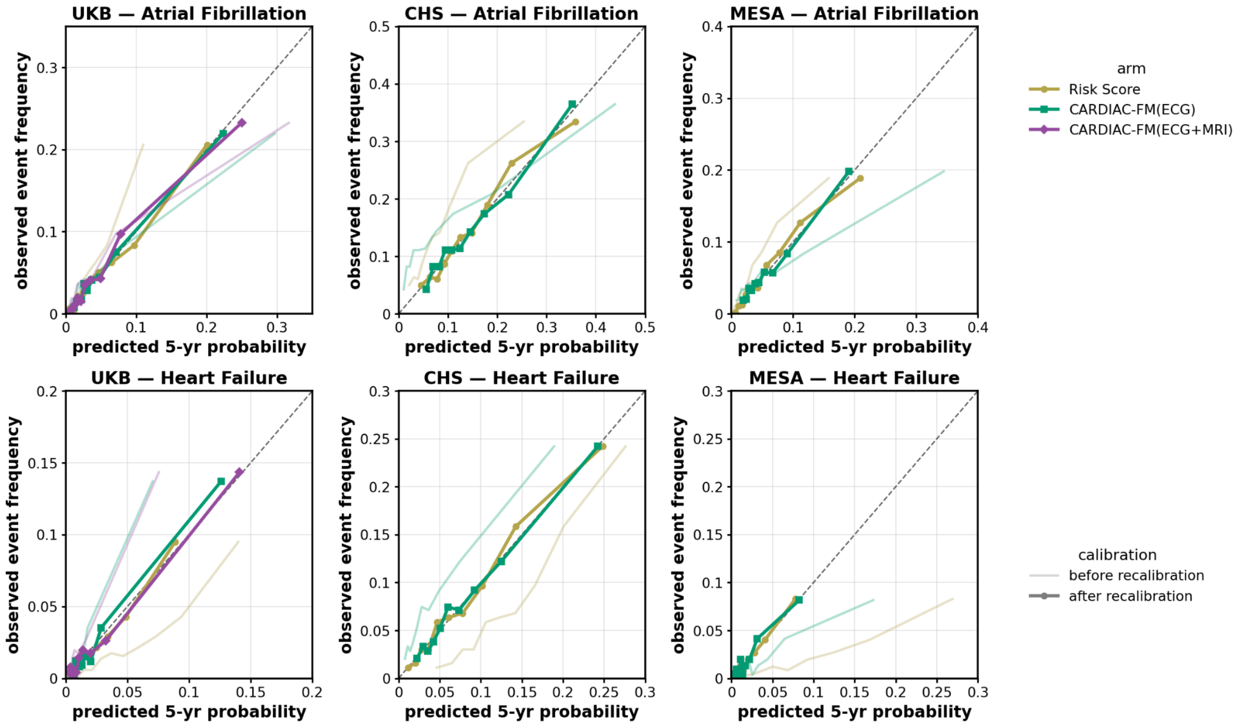


**Supplementary Figure 7 | Calibration plot for CARDIAC-FM and Risk Score.** Rows correspond to atrial fibrillation (top) and heart failure (bottom); columns correspond to the UK Biobank, CHS, and MESA cohorts. Predictions from each model (Risk Score, CARDIAC-FM with ECG only, and CARDIAC-FM with both ECG and MRI, where cardiac MRI was available) were grouped into deciles of predicted risk, and are compared against the observed event frequency. Each curve is shown before recalibration (faint lines) and after logistic recalibration (bold lines with markers). The diagonal dashed line indicates perfect calibration.

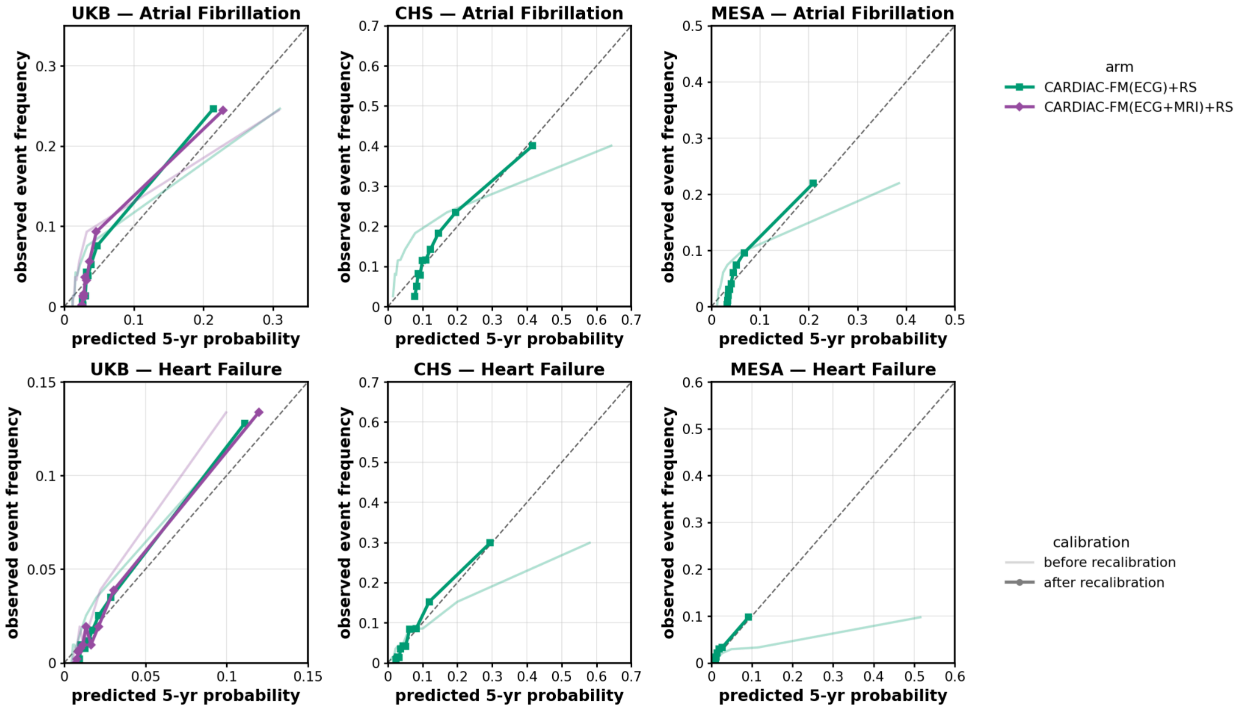


**Supplementary Figure 8 | Calibration plot for CARDIAC-FM combined with Risk Score.** Rows correspond to atrial fibrillation (top) and heart failure (bottom); columns correspond to the UK Biobank, CHS, and MESA cohorts. Predictions from each model (CARDIAC-FM with ECG only plus Risk Score, CARDIAC-FM with both ECG and MRI plus Risk Score, and CARDIAC-FM with both ECG and MRI, where cardiac MRI was available) were grouped into deciles of predicted risk, and are compared against the observed event frequency. Each curve is shown before recalibration (faint lines) and after logistic recalibration (bold lines with markers). The diagonal dashed line indicates perfect calibration.

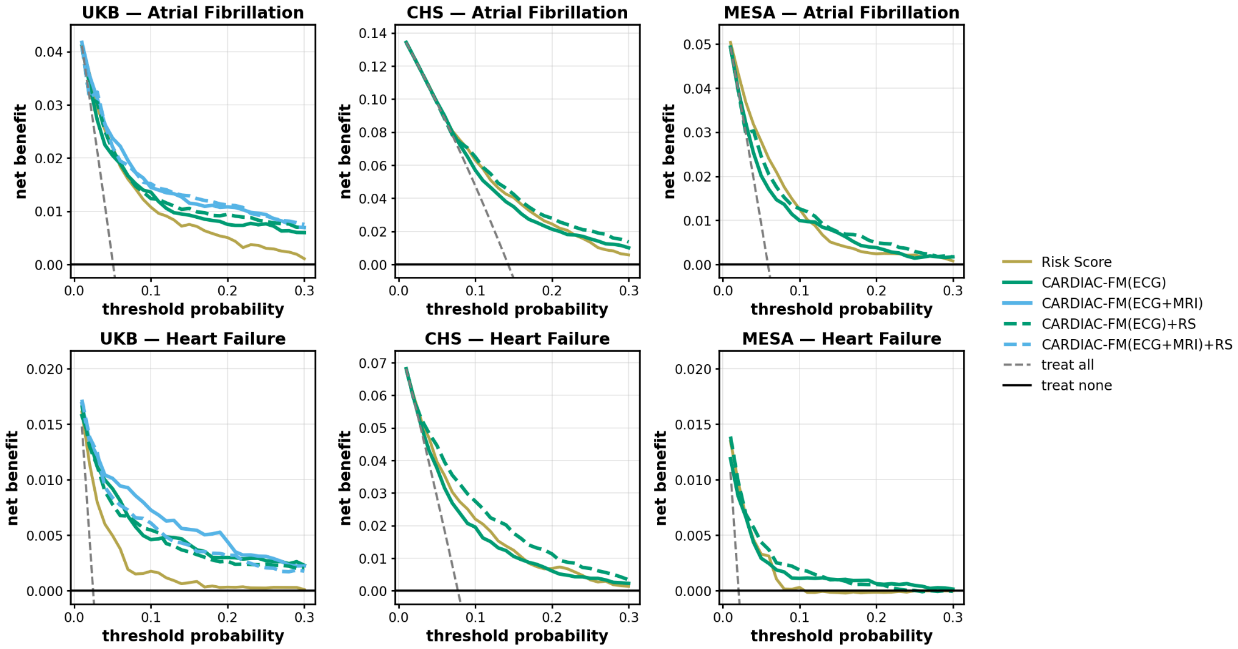


**Supplementary Figure 9 | Decision curves.** Rows correspond to atrial fibrillation (top) and heart failure (bottom); columns correspond to the UK Biobank, CHS, and MESA cohorts. Net benefit against the decision threshold probability for the clinical risk score, CARDIAC-FM (ECG), CARDIAC-FM (ECG + MRI), and CARDIAC-FM combined with the clinical risk score (CARDIAC-FM (ECG) + risk score and CARDIAC-FM (ECG + MRI) + risk score, when cardiac MRI was available). All arms were recalibrated by logistic scaling beforehand. The grey dashed line ("treat all") and the black solid line ("treat none") denote the reference strategies of predicting every participant as positive or negative, respectively.

(a) **Cardiovascular Health Study**

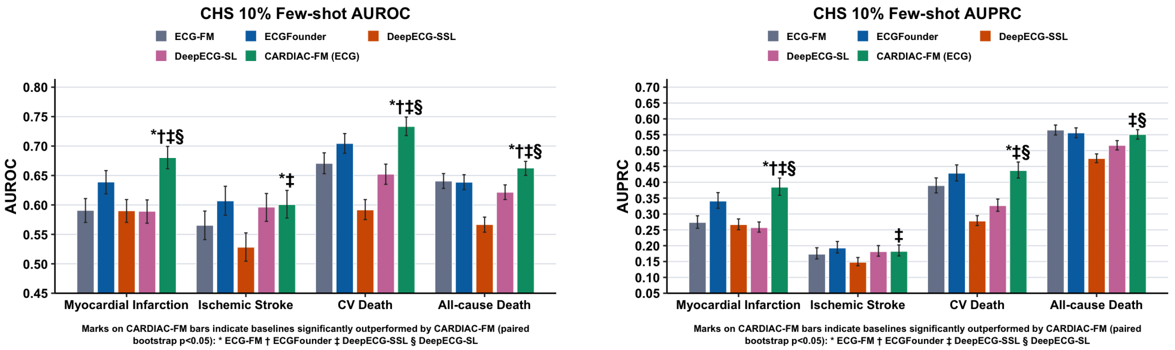


(b) **Multi-Ethnic Study of Atherosclerosis**

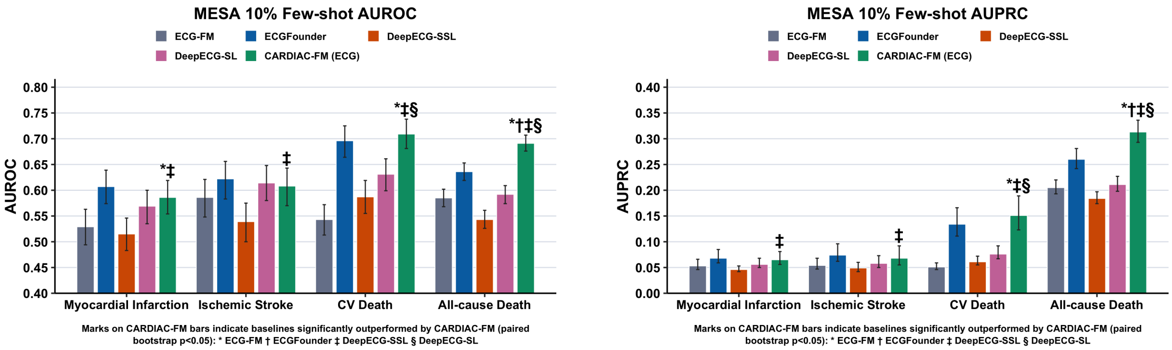


**Supplementary Figure 10 | Evaluation of pre-trained representations across a broad range of cardiovascular outcomes.** AUROC (left) and AUPRC (right) for CARDIAC-FM (blue) and ECG-FM (red) across four cardiovascular outcomes at a 10-year prediction horizon in CHS (**a**) and MESA (**b**). Outcomes include myocardial infarction (MI), ischaemic stroke (IS), cardiovascular death (CV death), and all-cause death (death). Both models used ECG-only input and were fine-tuned using 10% of data from each cohort. CARDIAC-FM consistently outperformed ECG-FM across all outcomes and time horizons in both cohorts. Error bars indicate 95% bootstrap confidence intervals.
